# Risk–benefit balance of habitual ultraviolet exposure for cardiovascular, cancer, and skin cancer mortality: UK Biobank cohort study

**DOI:** 10.64898/2026.01.08.26343592

**Authors:** Jiayue Gu, Andrew C. Stevenson, Annie R. Brady, Graeme J.M. Cowan, Chris Dibben, Richard B. Weller

**Author notes:** Corresponding authors: Jiayue. Gu, Institute for Regeneration and Repair, University of Edinburgh, 47 Little France Crescent EH16 4TJ, UK.

## Abstract

**Objective:** To examine how habitual ultraviolet (UV) exposure relates to cause-specific mortality and incidence, to quantify trade-offs between non-skin disease and skin cancer, and to explore potential circulating mediators.

**Design:** A population-based prospective cohort study with epidemiological and proteomic mediation analyses.

**Setting:** UK Biobank, recruited from 22 assessment centres across England, Scotland, and Wales.

**Participants:** 419 007 adults of White European ancestry with data on habitual UV exposure and follow-up for mortality and incident cardiovascular disease and cancer. A proteomic subcohort of 44 712 participants had plasma profiling.

**Main outcome measures:** Habitual ultraviolet exposure was summarised using Sun-BEEM (Sun-Behavioural and Environmental Exposure Model), a multidimensional score integrating environmental and behavioural indicators, categorised as low, medium, or high. Primary outcomes were all-cause, cardiovascular, and cancer mortality and incidence, and associations with Sun-BEEM categories were estimated using multivariable Cox models. Two extensions were implemented: an epidemiological extension using parametric g-computation to estimate deaths under counterfactual low and high UV scenarios; and a biological extension using proteomic mediation analyses to identify circulating proteins potentially linking UV exposure to cardiovascular and cancer mortality.

**Results:** Compared with low Sun-BEEM, medium and high exposure were associated with lower all-cause mortality (hazard ratio 0.89, 95% confidence interval 0.87 to 0.91; and 0.84, 0.82 to 0.87), with similar inverse associations for cardiovascular and non-skin cancer mortality. Skin cancer mortality showed no clear dose–response relationship with UV exposure, although incident keratinocyte cancers increased across Sun-BEEM categories. Counterfactual modelling suggested that, if associations are causal, a uniformly high UV pattern would prevent many more cardiovascular and other cancer deaths than the additional melanoma and keratinocyte cancer deaths. Proteomic mediation analyses implicated UV-downregulated immunoregulatory, mucosal–barrier, and cardiorenal–neuroendocrine pathways.

**Conclusions:** Higher habitual UV exposure, measured using a multidimensional score, was associated with lower cardiovascular and non-skin cancer mortality without clear increases in skin cancer mortality, supporting a more balanced view of sunlight and health.

**Summary box:** *What is already known on this topic:*

- Public health advice in temperate countries mainly treats sunlight as a skin cancer hazard.
- Few studies have explicitly quantified the trade-off between the potential benefits of habitual ultraviolet exposure for major non-skin diseases and its harms for skin cancer.
- Mechanistic research on how ultraviolet exposure affects health outcomes has focused largely on vitamin D, with only limited work on non–vitamin D pathways.

*What this study adds:*

- A multidimensional UV exposure score (Sun-BEEM), combining environmental and behavioural indicators, was associated with lower all-cause, cardiovascular, and non-skin cancer mortality, without clear increases in skin cancer mortality.
- Counterfactual analyses suggested a net balance favouring cardiovascular and cancer mortality benefits over skin cancer harms; proteomics supported mainly non–vitamin D pathways.

## Introduction

Cardiovascular disease and cancer are the leading causes of premature death in high income countries,[1, 2] and accumulating evidence suggests that sunlight exposure may influence the risk of both.[3,4] Yet public health messaging on sunlight is dominated by concern about skin cancer. In the UK, National Health Service (NHS) advice largely frames sunlight as a hazard, emphasising avoidance of strong sun (particularly between late morning and mid-afternoon), use of shade, clothing, and sunscreen, and recommending only limited exposure to maintain vitamin D status.[5,6] These recommendations were originally developed in high ultraviolet (UV) settings such as Australia,[7] where fair skinned populations live under subtropical conditions, and have been adopted in North-West Europe with little evaluation of their relevance to local UV levels, background risks, or competing causes of death[8]. By prioritising cutaneous risk and treating vitamin D as the main, if not sole, justification for intentional sun exposure, current guidance adopts a narrow view of sunlight that sits uneasily alongside emerging evidence for broader cardiometabolic and immunological effects of UV radiation. [4,9,10]

The epidemiology of sunlight and skin cancer further illustrates the gap between this risk-focused narrative and evidence on the wider health effects of UV exposure. UV radiation is a well-established cause of keratinocyte (non-melanoma) skin cancer, which occurs predominantly in the oldest age groups, but the relation to melanoma is less straightforward. [11–13] In many high-income countries, melanoma incidence has risen sharply while mortality has remained relatively stable,[14,15] a divergence widely attributed to increased scrutiny, diagnostic drift, and overdiagnosis, particularly of superficial spreading melanoma now classified as “low cumulative sun exposure” melanoma.[16–19] Much of the cohort literature has also relied on crude proxies of UV exposure, such as latitude, region of residence, or simple self-reported time outdoors,[20–22] and some widely cited estimates of sunlight-related skin cancer risk have been derived by comparing historical and contemporary cohorts—approaches that are highly sensitive to secular changes in diagnostic practice and health behaviours and are ill suited to quantifying absolute risks and benefits within a single population. On the benefit side, epidemiological and clinical discourse has often treated vitamin D as the main mediator of any health effects of sunlight, reinforcing the notion that oral supplementation can substitute for cutaneous UV exposure. Higher circulating 25-hydroxyvitamin D concentrations have been consistently associated with lower mortality from cardiovascular disease, cancer, melanoma, and diabetes, yet large vitamin D supplementation trials and Mendelian randomisation studies provide little evidence that vitamin D itself causally reduces these risks.[23–26] Experimental work has identified vitamin D–independent mechanisms by which UV exposure could influence systemic health, including photomobilisation of nitric oxide from cutaneous stores with downstream vascular effects[27,28] and immunoregulatory effects on antigen presenting cells and T cell responses,[29–31] suggesting that potentially beneficial systemic effects of habitual UV exposure remain under-recognised and insufficiently characterised at the population level.

Several cohort studies have related sunlight exposure to all-cause and cause-specific mortality, including non-skin outcomes using self-reported behaviours or area-based environmental indicators such as time spent outdoors, recent tanning history, skin colour, or region of residence.[32–34] However, these studies have typically focused on a limited set of endpoints and have rarely quantified the trade-offs between potential systemic benefits and skin cancer harms,[35] or identified circulating pathways that might underlie any associations. In the present study, we address these gaps using the Sun-BEEM score, a multidimensional measure of habitual UV exposure that combines long term residential UV climatology with individual-level behavioural information. We first examined the association between Sun-BEEM categories and all-cause, cardiovascular, cancer, and skin cancer mortality and incidence. For epidemiological extension, we then used parametric g-computation to estimate potential impact fractions and corresponding numbers of deaths under counterfactual low and high UV exposure scenarios,[36,37] aiming at quantifying the potential population impact of UV exposure to inform public health policy and to provide inputs for subsequent health economic and environmental modelling. For biological extension, we applied a prespecified proteomic mediation framework to identify circulating pathways that might mediate associations between habitual UV exposure and cause-specific mortality from cardiovascular disease and cancer.

## Methods

### Study population

This study used data from UK Biobank, a population-based prospective cohort of 502 492 participants aged 40-69 years recruited from 22 assessment centres across England, Scotland, and Wales between 2007 and 2010.[38] We used the most recent data release available as of 31 March 2025. At baseline, participants completed questionnaires on socioeconomic, lifestyle, and medical factors and provided blood and saliva samples. All gave written informed consent for long-term follow-up through linkage to National Health Service electronic health records. Analyses were restricted to participants of white European ancestry, using a combination of self-reported ethnic background and genetic information[39], to minimise confounding and population stratification related to differences in pigmentation/UV sensitivity, UV exposure patterns, and baseline risks of skin cancer and other health outcomes. Of 502 492 recruited participants, 453 026 with genetically inferred European ancestry were eligible, and 419 007 with data on habitual UV exposure and follow-up for health outcomes were included in the main analyses. Mediation analyses using plasma proteomic biomarkers were conducted in a subsample of 44 712 participants with Olink® Explore 1536 profiles (Olink Proteomics AB, Sweden), covering 2 494 proteins. Protein quantification and quality control followed standard UK Biobank/Olink pipelines (Supplementary Methods).

### Assessment of UV exposure: the Sun-BEEM score

Cumulative UV exposure was assessed using the Sun-BEEM (Behavioral and Environmental Exposure Model–UV) score, a composite index ranging from 0 to 4 that we developed specifically for the UK Biobank cohort. This represents the first implementation of a structured, multidimensional UV exposure model in UK Biobank, integrating both objective environmental data and self-reported UV-related behaviours. Sun-BEEM combines four domains of information: habitual time spent outdoors, residential ambient UV radiation derived from satellite data, self-reported solarium/sunlamp use, and use of sun/UV protection (figure 1).

**Figure 1.**
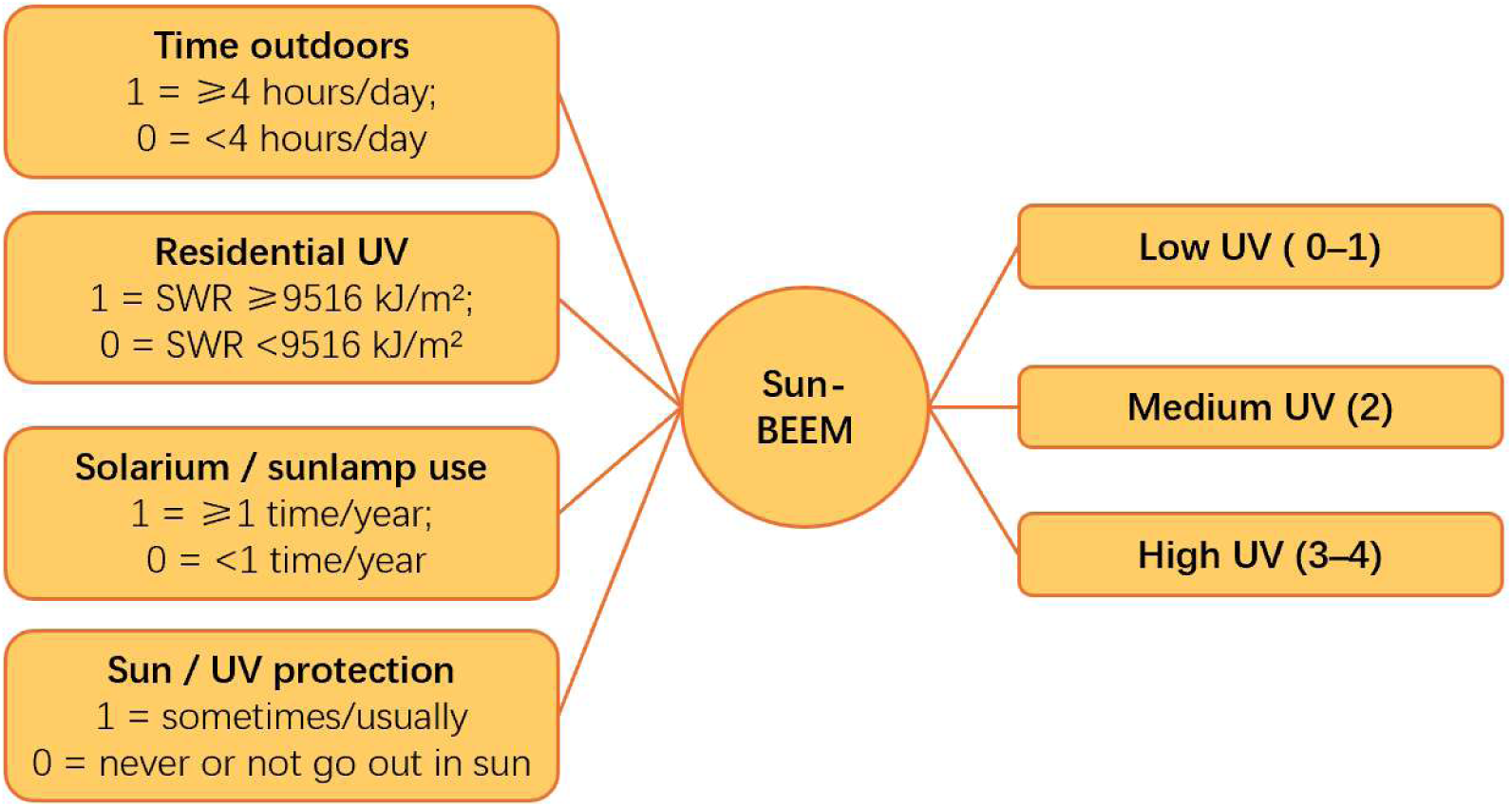
Construction of the Sun-BEEM (Behavioural and Environmental Exposure Model–UV) score. Sun-BEEM is a multidimensional index of habitual ultraviolet (UV) exposure derived from four binary components: time spent outdoors (1 = ≥4 hours/day; 0 = <4 hours/day, averaged across summer and winter), residential ambient UV (1 = annual downward shortwave radiation [SWR] ≥9516 kJ/m²; 0 = SWR <9516 kJ/m²), solarium/sunlamp use (1 = ≥1 time/year; 0 = <1 time/year), and sun/UV protection (1 = sometimes/usually uses protection; 0 = never/rarely uses protection or does not go out in the sun). Component scores are summed to give a total Sun-BEEM score from 0 to 4, which is then grouped into three exposure categories: low (0–1), medium (2), and high (3–4).

Habitual time spent outdoors was calculated from self-reported hours on a typical summer and winter day; the mean of these two values was dichotomised at 4 hours/day (score = 0 for <4 hours/day; score = 1 for ≥4 hours/day). Residential ambient solar radiation was estimated using the annual average downward shortwave radiation (SWR, kJ/m²) at each participant’s residential location, derived from data provided by the Japan Aerospace Exploration Agency. SWR was dichotomised at 9516 kJ/m², corresponding to the average annual value for Nottingham (<9516 kJ/m², score = 0; ≥9516 kJ/m², score = 1). Behavioural UV exposure was further characterised by solarium or sunlamp use (≥1 vs <1 time per year; score = 1 vs 0) and use of sun/UV protection (score = 1 for participants who sometimes used protection, and 0 for those who did not go out in sunshine or never used protection). All four binary components and their cut-offs were prespecified, based on behavioural and biological considerations, to capture meaningful contrasts in UV exposure. The four binary components were summed to yield a total Sun-BEEM score from 0 to 4, which was further grouped into low (0–1), medium (2), and high (3–4) UV exposure categories.

Each Sun-BEEM component was selected on the basis of prior evidence that it acts as a determinant or proxy of personal UV dose. [33,40–42] Details of variable derivation and cut-off selection are provided in the Supplementary Methods.

### Covariates and sensitivity analyses

We adjusted for potential confounders selected a priori based on biological plausibility and previous UK Biobank analyses. All main models were adjusted for age, sex, body mass index (BMI), Townsend Deprivation Index, highest educational attainment, smoking status, alcohol consumption, physical activity, and sleep quality. Age was modelled in categories, BMI using standard NHS categories, deprivation in quintiles of the Townsend score, and education in three levels. Smoking status, alcohol intake, physical activity, and sleep quality were modelled as categorical variables. Full definitions, categorisation procedure, and UK Biobank data-field identifiers are provided in the Supplementary Methods.

### Outcome ascertainment

Mortality outcomes were ascertained through linkage to national death registries (NHS Digital for England and Wales; NHS Central Register for Scotland), with follow-up to 31 March 2025. Underlying cause of death was coded using ICD-10. We examined all-cause mortality and cause specific mortality from cardiovascular disease (I21–I25, I60–I64), any cancer (C00–C97, D37–D48), melanoma (C43), other skin cancer (C44), and non-skin cancer (C00–C97, D37–D48 excluding C43 and C44).

Incident CVD events were identified from linked hospital inpatient records as the first hospital admission occurring after baseline with ICD-10 codes I21–I25 or I60–I64, among participants without a history of CVD prior to baseline. Incident cancers were ascertained from national cancer registries as the first cancer diagnosis after baseline, using ICD-10 site codes corresponding to the mortality definitions (overall cancer, melanoma, other skin cancer, and non-skin cancers), with participants with prevalent cancer at baseline excluded. Detailed ICD-10 definitions, outcome algorithms, and UK Biobank data-field identifiers are provided in the Supplementary Methods (Supplementary tables S4–S5).

### Statistical analysis

Baseline characteristics were summarised by Sun-BEEM exposure category (low [scores 0–1], medium [2], high [3–4]) to describe sociodemographic, lifestyle, and clinical profiles. Continuous variables were presented as means (standard deviations) or medians (interquartile ranges), and categorical variables as counts (percentages). Between-group differences were assessed using one-way analysis of variance for continuous variables and χ² tests for categorical variables. To assess construct validity of the Sun-BEEM score, baseline serum 25-hydroxyvitamin D [25(OH)D] concentrations were also compared across exposure categories. Although serum 25(OH)D was measured only once at baseline, within-person variation over time (for example due to season) is likely to be nondifferential and mean-zero, which would mainly increase standard errors and widen confidence intervals without biasing group means or their contrasts

To estimate associations between Sun-BEEM exposure and time-to-event outcomes, we fitted Cox proportional hazards models with time since baseline assessment as the underlying timescale. Time at risk was defined from baseline to the outcome or censoring, with follow-up censored on 31 March 2025, the latest date of complete mortality linkage; participants with incomplete or unreliable linkage were excluded. Primary analyses focused on mortality outcomes. For each endpoint, we fitted an unadjusted model and a multivariable model adjusted for age group, sex, body mass index, Townsend Deprivation Index, educational attainment, smoking status, alcohol intake, physical activity, and sleep quality (definitions in the Covariates section and Supplementary table S2). Missing data for covariates were addressed using multiple imputation by chained equations for sociodemographic and lifestyle variables, while occasional missing values for continuous variables were replaced with the sample mean. Sun-BEEM was modelled in three categories, with low exposure as the reference, and hazard ratios (HRs) with 95% confidence intervals (CIs) reported for medium and high versus low exposure. As a complementary analysis, incident cardiovascular and cancer events for the same disease categories were modelled using analogous cause specific Cox models, with censoring at competing causes or at the end of registry or hospital follow-up. Proportional hazards assumptions were assessed and confirmed using Schoenfeld residuals and log–log survival plots.

To assess the robustness of the main model, we undertook sensitivity analyses using alternative covariate specifications and study populations: (1) we fitted a series of nested Cox models for each mortality outcome, starting from a crude model and then sequentially adding sex, age, body mass index, deprivation, educational attainment, physical activity, sleep quality, smoking status, and alcohol intake; (2) landmark analyses excluding deaths within the first 2 and 5 years of follow-up; and (3) analyses stratified by sex, age group, and sunburn history. All sensitivity models used the same Cox framework and outcome definitions as the main multivariable model and are reported alongside the primary estimates.

For the epidemiological extension, parametric g-computation (the g-formula) was applied based on the main multivariable Cox mortality model under two counterfactual exposure scenarios. In a “low UV” scenario, all participants were reassigned to the Sun-BEEM low-exposure group; in a “high UV” scenario, all participants were reassigned to the high-exposure group. Using the fitted main model and holding covariates at their observed values, we first predicted, for each participant, the risk of each cause of death under the observed exposure distribution and under each counterfactual scenario and then summed these individual risks to obtain the corresponding expected numbers of deaths and mortality rates. For each cause of death (all-cause, cardiovascular, any cancer, melanoma, other skin cancer, and non-skin cancer), a potential impact fraction (PIF) was derived as the proportional difference between expected deaths under the observed and counterfactual exposure distributions. Positive PIFs indicate fewer expected deaths and lower mortality under the counterfactual (a protective shift), whereas negative PIFs indicate more deaths and higher mortality (a harmful shift). For each scenario, these relative impacts were then expressed in absolute terms by calculating the numbers of attributable deaths (fewer deaths) or excess deaths (additional deaths) as the difference between expected deaths under the observed and counterfactual exposure distributions.

For the biological extension, a two-stage mediation analysis was conducted in the proteomic subcohort to identify circulating biomarkers that might mediate associations between UV exposure and cause-specific mortality from cardiovascular disease and cancer. This analysis used the same mortality outcomes, Sun-BEEM exposure parameterisation, and covariate set as the main multivariable Cox mortality model. In the first stage, for each biomarker we fitted two Cox models for the outcome: (i) a model including Sun-BEEM and covariates (total effect) and (ii) a model additionally including the biomarker (direct effect). The difference between the Sun-BEEM coefficients on the log-HR scale was interpreted as the indirect effect, and the proportion mediated was calculated as (total effect − direct effect) / total effect. Ninety-five per cent CIs for the indirect effect were obtained using non-parametric bootstrap resampling with 1,000 iterations. Biomarkers with sparse data (fewer than 100 participants with valid measurements) or unstable bootstrap estimates (fewer than 10 successful bootstrap replications) were excluded. In the second stage, associations between Sun-BEEM and each biomarker were examined using multivariable linear regression, adjusted for the same covariates as the main mortality model. Only biomarkers with sufficient data coverage (≥100 participants with non-missing biomarker values) were retained. P values for the UV–biomarker associations were adjusted for multiple testing using the Benjamini–Hochberg false discovery rate (FDR) procedure. Biomarkers were considered putative mediators if they met both of the following criteria: (1) the bootstrapped 95% CI for the indirect effect excluded zero; and (2) the FDR-adjusted P value for the UV–biomarker association was <0.01. Because biomarkers were evaluated in separate mediation models and many map to overlapping biological pathways, the resulting mediation proportions were used primarily to screen and rank candidate mediators, rather than to estimate mutually independent or additive pathway-specific effects.

## Results

### Population characteristics and validation of the Sun-BEEM score

Among 419 007 participants included in the main analyses, just over half (53.5%) were women and the mean age was 56.1 years (SD 8.0). Baseline characteristics varied across Sun-BEEM categories (table 1). Compared with the low UV group, participants in the high UV group were more often men (52% v 46%), more likely to report high physical activity (46% v 27%), and slightly less likely to be obese (23% v 25%). The proportion living in the most deprived quintile was slightly higher in the high Sun-BEEM group than in the low group, but absolute differences were small; those in the high UV group were less likely to hold a university degree (40% v 49%). Current or former smoking (52% v 44%) and higher alcohol intake (43% v 39%) were also more frequent in the high UV group, whereas sleep scores were similar across categories. Reported childhood sunburn episodes were infrequent and showed no clear gradient across Sun-BEEM categories, with mean values ranging from about 1.6 to 1.9 episodes.

**Table 1.**
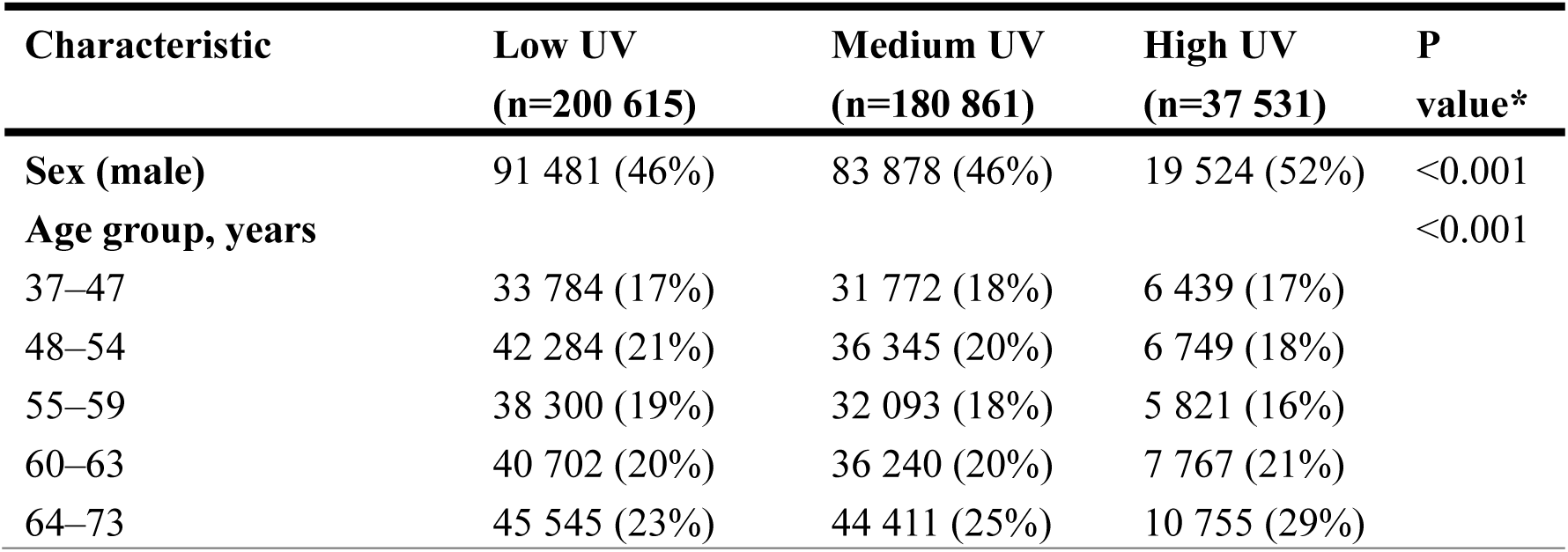
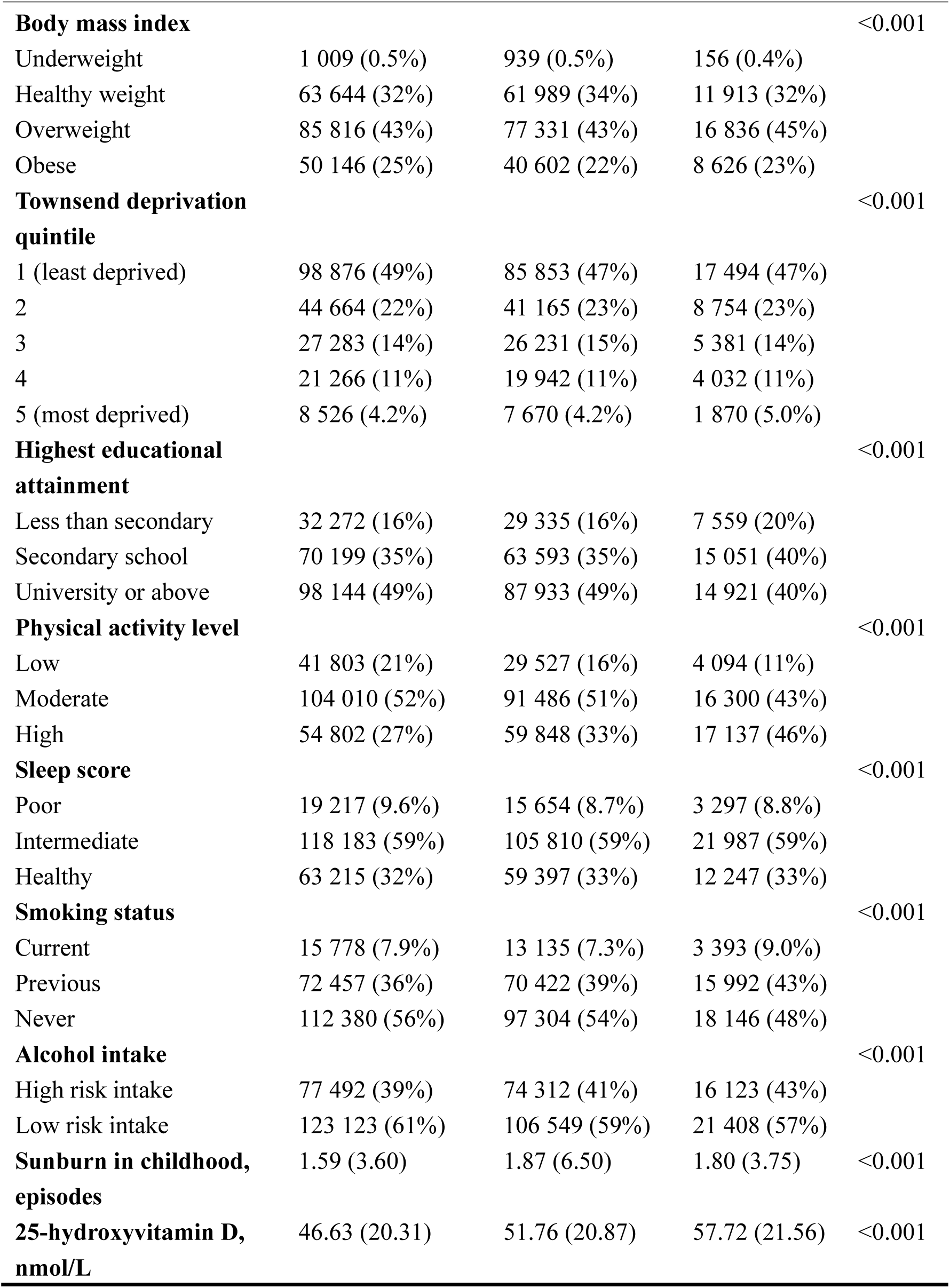
Baseline characteristics of participants by Sun-BEEM ultraviolet exposure category. Values are n (%) unless stated otherwise; continuous variables are mean (standard deviation). P values are for differences across Sun-BEEM categories (χ² tests for categorical variables and analysis of variance for continuous variables). Categories for body mass index, physical activity level, sleep score, smoking status and alcohol intake are defined in supplementary methods.

| Characteristic | Low UV<br>(n=200 615) | Medium UV<br>(n=180 861) | High UV<br>(n=37 531) | P<br>value* |
| --- | --- | --- | --- | --- |
| **Sex (male)** | 91 481 (46%) | 83 878 (46%) | 19 524 (52%) | $<0.001$ |
| **Age group, years** | | | | $<0.001$ |
| 37–47 | 33 784 (17%) | 31 772 (18%) | 6 439 (17%) |  |
| 48–54 | 42 284 (21%) | 36 345 (20%) | 6 749 (18%) |  |
| 55–59 | 38 300 (19%) | 32 093 (18%) | 5 821 (16%) |  |
| 60–63 | 40 702 (20%) | 36 240 (20%) | 7 767 (21%) |  |
| 64–73 | 45 545 (23%) | 44 411 (25%) | 10 755 (29%) |  |
| **Body mass index** |  |  |  | <0.001 |
| Underweight | 1 009 (0.5%) | 939 (0.5%) | 156 (0.4%) |  |
| Healthy weight | 63 644 (32%) | 61 989 (34%) | 11 913 (32%) |  |
| Overweight | 85 816 (43%) | 77 331 (43%) | 16 836 (45%) |  |
| Obese | 50 146 (25%) | 40 602 (22%) | 8 626 (23%) |  |
| **Townsend deprivation quintile** |  |  |  | <0.001 |
| 1 (least deprived) | 98 876 (49%) | 85 853 (47%) | 17 494 (47%) |  |
| 2 | 44 664 (22%) | 41 165 (23%) | 8 754 (23%) |  |
| 3 | 27 283 (14%) | 26 231 (15%) | 5 381 (14%) |  |
| 4 | 21 266 (11%) | 19 942 (11%) | 4 032 (11%) |  |
| 5 (most deprived) | 8 526 (4.2%) | 7 670 (4.2%) | 1 870 (5.0%) |  |
| **Highest educational attainment** |  |  |  | <0.001 |
| Less than secondary | 32 272 (16%) | 29 335 (16%) | 7 559 (20%) |  |
| Secondary school | 70 199 (35%) | 63 593 (35%) | 15 051 (40%) |  |
| University or above | 98 144 (49%) | 87 933 (49%) | 14 921 (40%) |  |
| **Physical activity level** |  |  |  | <0.001 |
| Low | 41 803 (21%) | 29 527 (16%) | 4 094 (11%) |  |
| Moderate | 104 010 (52%) | 91 486 (51%) | 16 300 (43%) |  |
| High | 54 802 (27%) | 59 848 (33%) | 17 137 (46%) |  |
| **Sleep score** |  |  |  | <0.001 |
| Poor | 19 217 (9.6%) | 15 654 (8.7%) | 3 297 (8.8%) |  |
| Intermediate | 118 183 (59%) | 105 810 (59%) | 21 987 (59%) |  |
| Healthy | 63 215 (32%) | 59 397 (33%) | 12 247 (33%) |  |
| **Smoking status** |  |  |  | <0.001 |
| Current | 15 778 (7.9%) | 13 135 (7.3%) | 3 393 (9.0%) |  |
| Previous | 72 457 (36%) | 70 422 (39%) | 15 992 (43%) |  |
| Never | 112 380 (56%) | 97 304 (54%) | 18 146 (48%) |  |
| **Alcohol intake** |  |  |  | <0.001 |
| High risk intake | 77 492 (39%) | 74 312 (41%) | 16 123 (43%) |  |
| Low risk intake | 123 123 (61%) | 106 549 (59%) | 21 408 (57%) |  |
| **Sunburn in childhood, episodes** | 1.59 (3.60) | 1.87 (6.50) | 1.80 (3.75) | <0.001 |
| **25-hydroxyvitamin D, nmol/L** | 46.63 (20.31) | 51.76 (20.87) | 57.72 (21.56) | <0.001 |

Sun-BEEM showed a clear gradient with serum 25-hydroxyvitamin D [25(OH)D], with mean concentrations rising from 46.63 nmol/L in the low group to 51.76 nmol/L in the medium group and 57.72 nmol/L in the high group (p<0.001). In fully adjusted linear regression, each one-category increase in Sun-BEEM was associated with a 4.46 nmol/L higher 25(OH)D (95% confidence interval 4.32 to 4.59; p<2×10⁻¹⁶), supporting its use as a proxy for cumulative UV exposure in this cohort.

### Associations of habitual UV exposure with mortality and incidence

In fully adjusted Cox models accounting for age, sex, body mass index, deprivation, education, smoking, alcohol intake, physical activity, and sleep quality, higher Sun-BEEM exposure was associated with lower risks of several major mortality outcomes compared with the low exposure group (figure 2A). Medium and high UV exposure were both associated with reduced all-cause mortality (hazard ratio 0.89, 95% confidence interval 0.87 to 0.91; and 0.84, 0.82 to 0.87, respectively), with strong evidence of a dose–response trend (p<0.001). Similar inverse associations were seen for cardiovascular mortality (0.82, 0.78 to 0.87 for medium; 0.77, 0.71 to 0.85 for high) and for non-skin cancer mortality (0.92, 0.89 to 0.94 for medium; 0.89, 0.85 to 0.93 for high).

**Figure 2.**
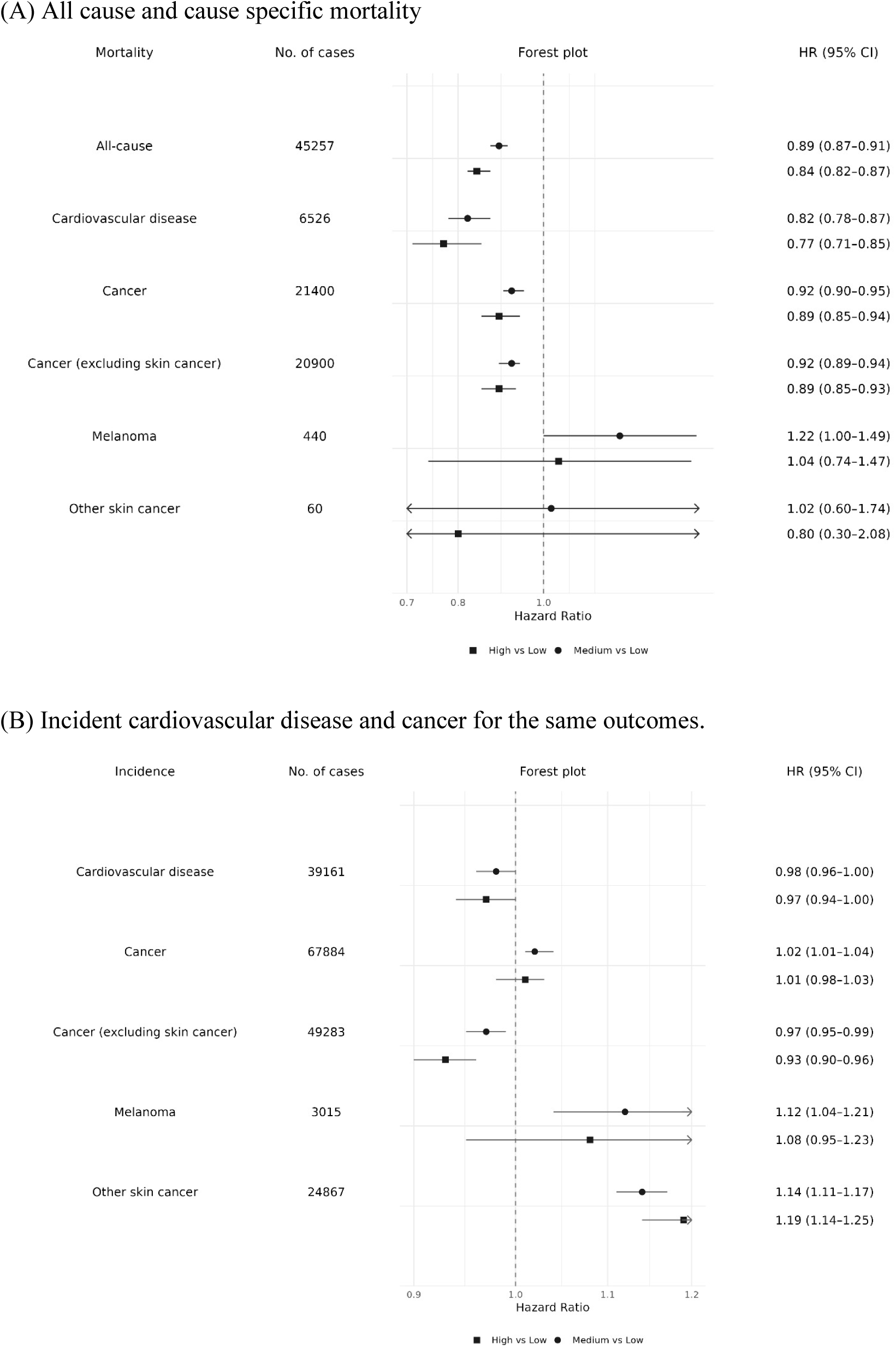
Associations of habitual ultraviolet exposure with disease mortality and incidence (A) All cause and cause specific mortality. (B) Incident cardiovascular disease and cancer for the same outcomes. Low Sun-BEEM exposure is the reference category; medium (circle) and high exposure (square) categories are compared with this group. Horizontal lines indicate 95% confidence intervals.

No clear associations were observed for skin cancer mortality. Among 440 melanoma deaths, there was a borderline increase in risk in the medium UV group (1.22, 1.00 to 1.49) but no convincing association in the high UV group (1.04, 0.74 to 1.47). Mortality from other skin cancers was imprecisely estimated, with wide confidence intervals (1.02, 0.60 to 1.74 for medium; 0.80, 0.30 to 2.08 for high), reflecting the small number of deaths. Proportional hazards assumptions for Sun-BEEM were not materially violated in the mortality models.

Complementary analyses of incident events (figure 2B) showed broadly consistent patterns. Higher UV exposure was associated with modest reductions in incident cardiovascular disease (0.98, 0.96 to 1.00 for medium; 0.97, 0.94 to 1.00 for high) and in incident cancer excluding skin cancers (0.97, 0.95 to 0.99 for medium; 0.93, 0.90 to 0.96 for high), with confidence intervals for cardiovascular outcomes close to the null. By contrast, incident melanoma and other skin cancers increased with higher Sun-BEEM categories (melanoma: 1.12, 1.04 to 1.21 for medium; 1.08, 0.95 to 1.23 for high; other skin cancer: 1.14, 1.11 to 1.17 for medium; 1.19, 1.14 to 1.25 for high). Overall, these findings indicate a consistent inverse association between habitual UV exposure and cardiovascular and non-skin cancer outcomes. Moderate, but not high sun exposure was associated with increased incident melanoma. ‘Other skin cancer’ which is a composite measure made up predominantly of basal cell cancers and squamous cell skin cancers showed a biological gradient of increased incidence with increased sun exposure.

Findings were broadly similar across the three sets of sensitivity analyses (supplementary tables S1–S6). In sequentially adjusted (nested) Cox models, most attenuation from the crude estimates occurred after adjustment for sex and age, whereas further adjustment for body mass index, deprivation, educational attainment, physical activity, sleep quality, smoking status, and alcohol intake led to only small additional changes and did not alter the overall pattern of associations. Landmark analyses excluding deaths within the first 2 and 5 years of follow-up produced effect estimates of similar magnitude to the main analyses, suggesting that reverse causation is unlikely to fully explain the observed associations. Stratified analyses by sex, age group, and sunburn history also showed broadly consistent patterns, with no strong evidence of effect modification.

### Population impact of counterfactual UV exposure scenarios

Using parametric g-computation based on the main multivariable Cox mortality model, we estimated potential impact fractions (PIFs) for two counterfactual exposure scenarios in which all participants were reassigned to a single Sun-BEEM category. In the low UV counterfactual scenario, where the entire population was reassigned to the Sun-BEEM low exposure group, PIFs were positive for melanoma mortality (PIF 9.0%) and keratinocyte cancer mortality (PIF 0.9%), corresponding to an estimated 39 and 1 fewer deaths, respectively, compared with the observed exposure distribution. By contrast, PIFs were negative for all-cause mortality (–6.6%), cardiovascular mortality (–10.9%), and all cancer mortality (–4.6%), corresponding to an estimated 2 982,711, and 994 additional deaths, respectively (figure 3A and 3B).

**Figure 3.**
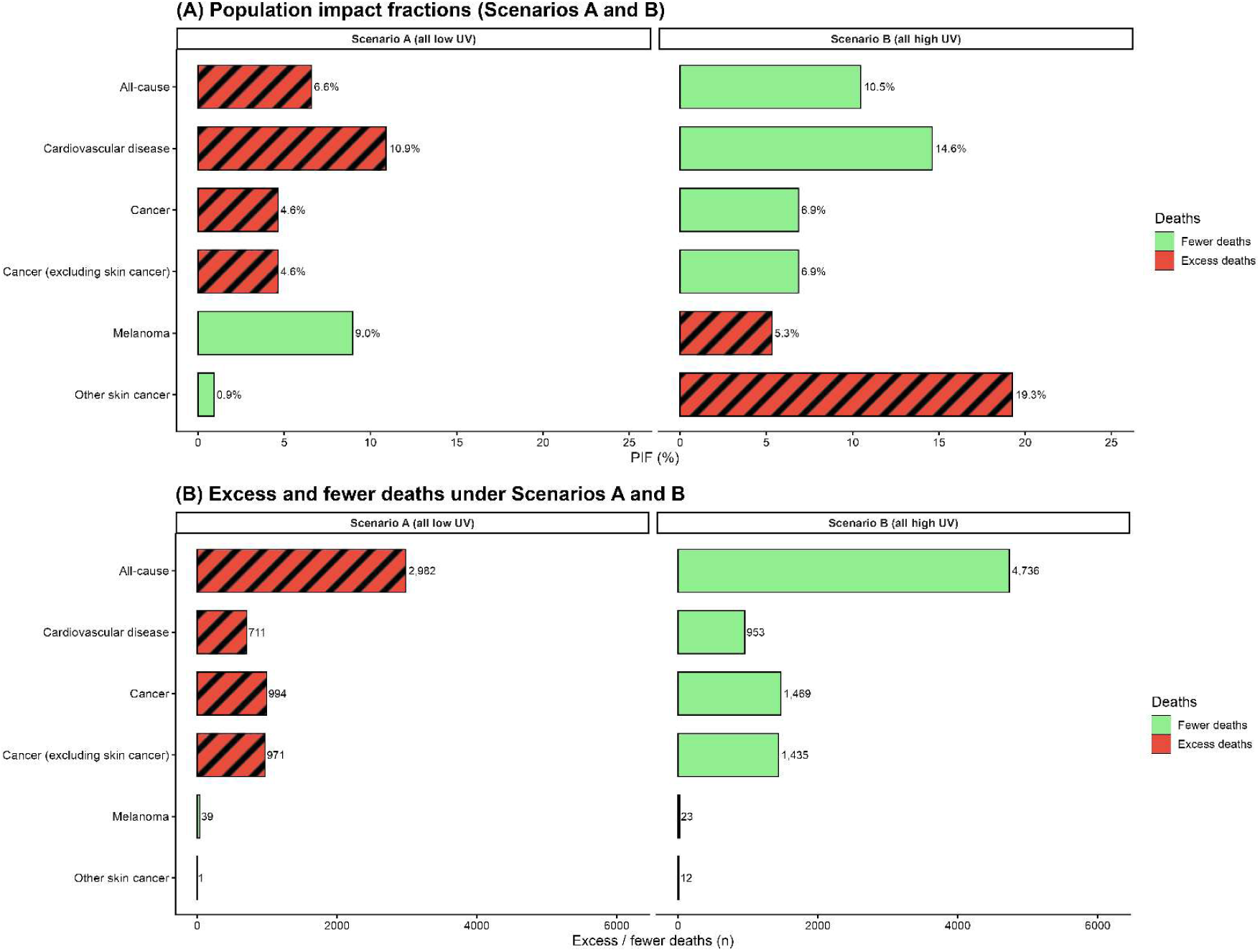
Population impact of ultraviolet (UV) exposure on mortality under two scenarios (A) Population impact fractions (PIFs) for all outcomes under counterfactual scenarios in which all participants are reassigned to low UV exposure (Scenario A) or high UV exposure (Scenario B); solid bars indicate positive PIF values and striped bars indicate negative PIF values. (B) Corresponding numbers of fewer or excess deaths relative to the observed exposure distribution, with solid bars indicating fewer deaths and striped bars indicating excess deaths.

In the high UV counterfactual scenario, where all participants were reassigned to the Sun-BEEM high exposure group, PIFs were positive for all-cause mortality (10.5%), cardiovascular mortality (14.6%), and all cancer mortality (6.9%), corresponding to an estimated 4 736, 953, and 1 469 fewer deaths, respectively. PIFs were negative for keratinocyte cancer mortality (–19.3%) and melanoma mortality (–5.3%), corresponding to an estimated 12 and 23 additional deaths, respectively (figure 3A and 3B). On balance, these counterfactual scenarios suggest that a uniformly low UV exposure pattern would be dominated by excess cardiovascular and all-cause deaths, whereas a uniformly high exposure pattern would be dominated by fewer deaths in these outcomes at the cost of some excess deaths from skin cancers.

### Proteomic mediation of associations between UV and mortality

To explore biological mechanisms that might link UV exposure to mortality, we conducted mediation analysis in the proteomic sub-cohort (figure 4). This analysis identified circulating proteins that met prespecified criteria for potentially mediating the association between Sun-BEEM exposure and cardiovascular or cancer mortality.

**Figure 4.**
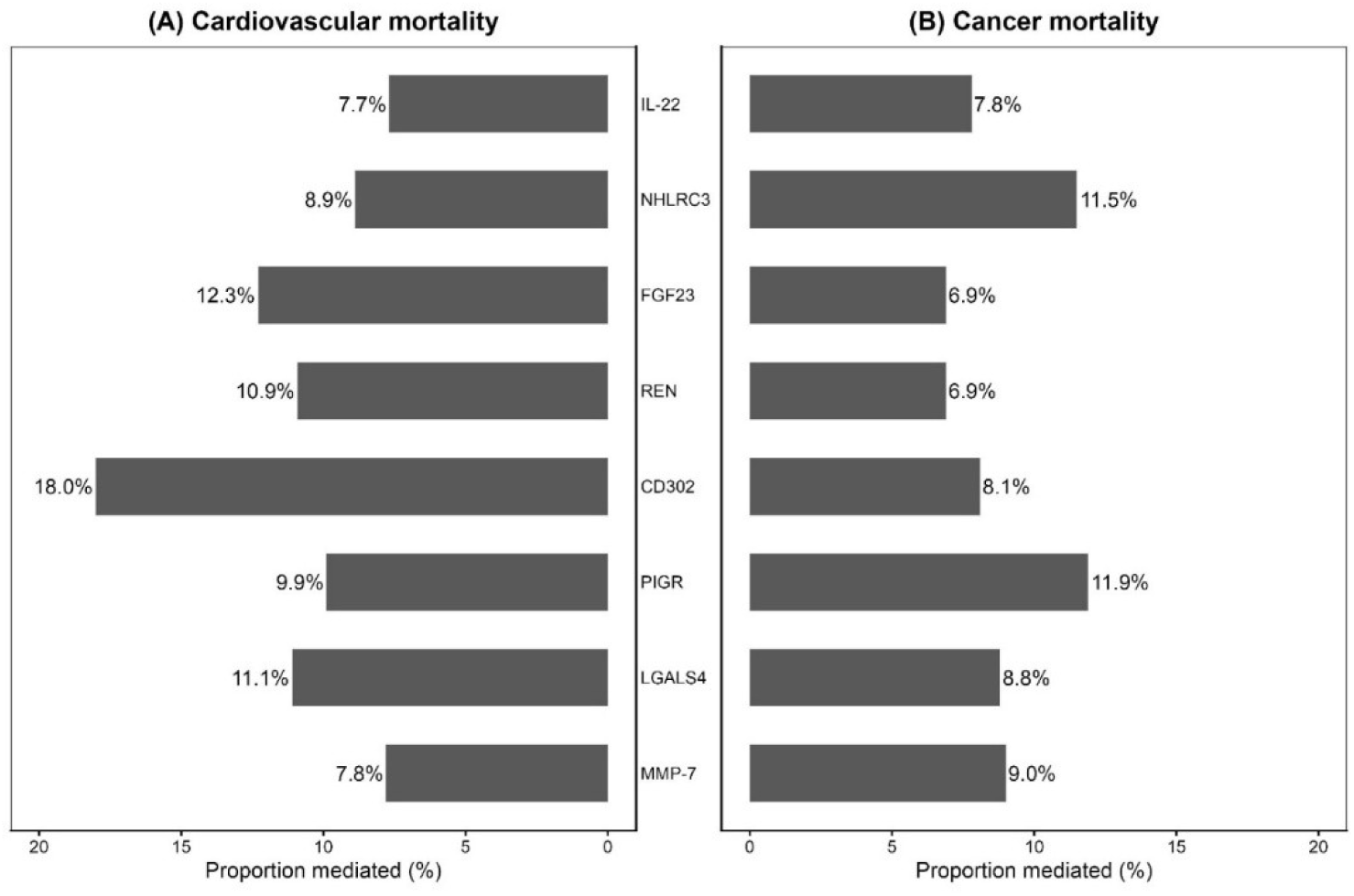
Overlapping proteins mediating the association between UV exposure and mortality. Bars show the proportion of the total association mediated by each protein for cardiovascular (A) and cancer (B) mortality. Eight biomarkers (IL-22, NHLRC3, FGF23, REN, CD302, PIGR, LGALS4, and MMP-7) overlapped as putative mediators of both outcomes.

Across outcomes, eight biomarkers overlapped as putative mediators of both cardiovascular and cancer mortality: PIGR, NHLRC3, MMP-7, CD302, FGF23, REN, LGALS4, and IL-22 (figure 4; supplementary table S11). For cardiovascular mortality, these proteins each mediated between 7.7% and 18.0% of the total UV effect, with CD302 showing the largest proportion (18.0%; figure 4A). For cancer mortality, the same eight proteins mediated between 6.9% and 11.9% of the total effect, with PIGR and NHLRC3 among the strongest mediators (11.9% and 11.5%, respectively; figure 4B). Full ranked lists of proteins mediating the associations with cardiovascular and cancer mortality are shown in supplementary table S9-S10. Because many of these markers are biologically correlated and act within overlapping pathways, the reported mediation percentages are best interpreted as relative indicators of mediating influence, rather than as precise, independent shares of the total UV–mortality association that can be directly compared or summed across proteins.

## Discussion

### Interpretation of mortality and incidence associations with UV exposure

In this large prospective analysis of 419,007 UK Biobank participants, higher habitual UV exposure—quantified using the multidimensional Sun-BEEM index—was associated with lower all-cause, cardiovascular, and non-skin cancer mortality in a dose-dependent fashion. Compared with those in the lowest Sun-BEEM category, participants in the highest category had lower risks of death from cardiovascular disease and internal cancers, and these associations were robust to extensive adjustment for sociodemographic, behavioural, and clinical factors and to multiple prespecified sensitivity analyses.

Our findings sit within, and extend, a small but provocative literature on sunlight and internal-cause mortality. In the Melanoma in Southern Sweden (MISS) cohort, avoidance of sun exposure was associated with roughly a doubling of all-cause mortality compared with the highest sun-exposure group, largely because of more deaths from cardiovascular and non-cancer causes, suggesting that very low sun exposure may carry independent health risks.[43] Taken together with our results, this indicates a broadly consistent pattern in which higher habitual solar UV exposure is associated with lower all-cause, cardiovascular, and non-skin cancer mortality. By contrast, studies reporting higher mortality in more “sun-damaged” individuals have often relied on surrogate measures such as dermatologist-rated actinic skin damage (all-cause, cardiovascular, and cancer mortality in the National Health and Nutrition Examination Survey (NHANES I),[44] composite “visible ageing” signs including baldness, earlobe crease, and xanthelasmata (incident ischaemic heart disease and myocardial infarction),[45] or deep forehead wrinkles (all-cause and cardiovascular mortality in a working cohort)[46]—indices that strongly reflect age, smoking, skin type, and comorbidities and are therefore difficult to interpret as specific, causal measures of UV dose in relation to internal-cause mortality.

Although UV radiation is a recognised cause of skin cancers, we did not observe a strong positive association between higher habitual UV exposure in middle age and skin cancer mortality. Melanoma mortality was modestly higher in the intermediate Sun-BEEM category (HR≈1.22; 95% CI 1.00–1.49) but not in the highest category (HR≈1.04; 0.74–1.47), and estimates were imprecise because of the small number of deaths and could be compatible with chance. This non-linear pattern may reflect threshold or adaptive responses to chronic UV exposure—such as epidermal thickening, increased pigmentation, and UV-induced immunomodulation—so that risk does not increase in a simple dose–response fashion with cumulative dose.[29] The timing of exposure is also likely to be critical: childhood and early-adult intermittent “holiday” sun exposure promotes melanocytic naevus development on intermittently exposed sites, and naevus burden is one of the strongest phenotypic predictors of melanoma risk,[47] whereas UK Biobank recruited participants aged 40 years and older and thus captures habitual midlife exposure rather than this early-life susceptibility window. Against this background, our data suggest that, within the exposure range and age span captured in this cohort, increased midlife UV exposure in this UK population is unlikely to confer a large increase in fatal melanoma risk, although modest effects in either direction cannot be excluded. For non-melanoma skin cancer, despite substantial UV-related morbidity and a large share of cancer-related healthcare encounters, mortality in the UK is very low—accounting for around 1% of all cancer deaths—and heavily concentrated at older ages, with about three quarters (76%) of non-melanoma skin cancer deaths occurring in people aged 75 years and over and mortality rates highest in those aged 90 years and above.[48] Because follow-up ended when participants had reached a mean attained age of approximately 73 years, and relatively few individuals reached very old age during follow-up, any excess risk of keratinocyte cancer deaths emerging predominantly in very old age would be difficult to detect, which may further contribute to the absence of a clear gradient between Sun-BEEM and non-melanoma skin cancer mortality. Within the same Cox modelling framework, therefore, higher habitual UV exposure was associated with appreciable reductions in cardiovascular and other internal-cause mortality, but no clear increase in deaths from melanoma or other skin cancers.

Analyses of incident events broadly supported these mortality findings while adding nuance. For cancers other than skin cancer, higher Sun-BEEM categories were associated with lower incidence, mirroring the dose–response pattern seen for mortality. Because these outcomes were ascertained independently through national cancer registrations, this concordance suggests it is unlikely that the inverse associations for internal cancers are explained solely by misclassification or selective survival. By contrast, associations with incident cardiovascular disease were weaker and not statistically distinguishable from unity, with confidence intervals compatible with little or no effect. This may reflect under-ascertainment of less severe or non-hospitalised events, but it is also plausible that habitual UV exposure has a greater impact on disease progression and fatal complications than on the initial development of cardiovascular disease. In addition, associations for incident outcomes may have been attenuated by selection effects, because participants with relevant pre-baseline hospital admissions were excluded, leaving a healthier cohort for incident analyses compared with mortality. For melanoma, both incidence and mortality were slightly higher in the higher Sun-BEEM categories, but there was no clear dose–response gradient and confidence intervals were compatible with at most modest effects; these patterns are consistent with the complex relation between UV exposure and melanoma risk and should be interpreted with caution. For other skin cancers, the number of deaths was too small to allow reliable estimation of mortality associations, but there were several tens of thousands of incident cases, and these exhibited a clear dose–response increase in incidence across Sun-BEEM categories, in keeping with UV radiation as a primary carcinogen for keratinocyte cancers; this pattern supports the construct validity of our exposure metric and argues against major systematic bias favouring protective associations for internal disease outcomes.

### Population-level impact and trade-offs across causes of death

In counterfactual analyses using parametric g-computation, we explored what these associations might imply at a population level. When we modelled a scenario in which everyone in the cohort had high Sun-BEEM exposure, we estimated that, if the associations are causal, around 5 000 deaths during follow-up might have been avoided, largely because of fewer deaths from cardiovascular disease and cancers other than skin cancer, in exchange for only a few dozen additional deaths from melanoma and keratinocyte cancers combined. Conversely, a scenario in which everyone had low Sun-BEEM exposure produced the opposite pattern: only a few dozen fewer skin cancer deaths but around 3 000 additional deaths from cardiovascular disease and other cancers. These scenario-based estimates are intended to illustrate the potential population impact of different UV exposure patterns rather than to provide precise forecasts, and they depend on the causal interpretation of the underlying associations.

To our knowledge, no previous work has quantified these trade-offs within a single analytical framework. Earlier cohort studies have reported apparently protective associations between greater sun exposure and cardiovascular or all-cause mortality but have not examined how many deaths would be gained or lost under alternative exposure patterns. By contrast, a substantial literature—and much public messaging—has focused on attributing melanoma incidence to UV exposure, often quoting statements that “around 85 out of 100 melanomas are caused by too much ultraviolet (UV) radiation”.[49] These figures do not arise from measured individual UV exposure, but from incidence-based population attributable fractions that designate contemporary birth cohorts as “high UV–exposed” and early twentieth century (Edwardian) cohorts as “low UV–exposed” on the assumption that the latter “almost certainly had little bodily exposure to sunlight” in childhood.[50] In effect, year of birth is used as a proxy for UV dose, with only limited capacity to account for concurrent changes in clothing, indoor lifestyles, occupational structure, competing risks, or, critically, the intensity of dermatological surveillance and biopsy. By directly modelling individual habitual UV exposure with the multidimensional Sun-BEEM index, the present study avoids treating birth cohort as a surrogate for UV dose and provides a more appropriate basis for quantifying both the harms and the benefits of UV exposure. On balance, these considerations not only introduce an explicit trade-off perspective into the epidemiology of UV exposure but also support a broader reappraisal of melanoma epidemiology. [51–52] Recent analyses of UK melanoma mortality, including our own work, have suggested that UV radiation is unlikely to be the predominant driver of fatal melanoma in contemporary high-surveillance settings [53–54]; the present counterfactual analyses build on this by suggesting that, at the population level, modest increases in skin cancer deaths associated with higher UV exposure would, if causal, be outweighed by larger reductions in deaths from cardiovascular disease and other cancers.

These findings together suggest that an important public health question is how different patterns of UV exposure redistribute deaths across causes. Over and above this redistribution, it also matters when those deaths occur and what life is like beforehand: compared with skin cancers, cardiovascular disease and other major internal cancers are more likely to cause deaths at younger ages and to be preceded by sustained impairments in health-related quality of life. National statistics are consistent with this pattern: in the UK, cardiovascular disease accounts for around 49 000 deaths each year in people younger than 75 years and roughly a quarter of all premature deaths, [55–56] whereas more than half of melanoma deaths and about three quarters of non-melanoma skin cancer deaths occur in those aged 75 years and over, with mortality rates for both cancers highest among people aged 90 years and above. [48,57]

On average, cardiovascular disease and major internal cancers remove more years of good-quality life than melanoma and, especially, keratinocyte cancers. Cohort and health-economic studies typically report EQ-5D utilities in the range of about 0.6–0.8 among survivors of stroke or myocardial infarction in the years after the event, on a scale where 1.0 represents full health and 0 represents death.[58,59] In contrast, utilities for advanced melanoma used in NICE technology appraisals fall markedly in the months before death,[60] and most non-melanoma skin cancers and actinic keratoses are associated with EQ-5D utilities clustering between about 0.85 and 0.95, close to age-matched population norms. [61,62] These patterns suggest that, if the associations we observe are causal, higher habitual UV exposure would preserve many more years of good-quality life by reducing deaths from cardiovascular disease and other major internal cancers.

### Potential biological mechanisms underlying the observed associations

To our knowledge, this is the first large prospective cohort study to relate habitual UV exposure to cardiovascular and cancer mortality using a formal proteomic mediation framework, explicitly linking exposure, circulating proteins, and subsequent disease outcomes. In mediation analyses, eight UV-downregulated proteins were grouped on functional grounds into three axes—immunoregulatory (IL-22 and NHLRC3), mucosal–barrier/innate (MMP-7, CD302, PIGR, and LGALS4), and cardiorenal–neuroendocrine (FGF23 and REN)—spanning both cardiovascular and cancer mortality.

On the immunoregulatory axis, higher Sun-BEEM exposure was associated with lower circulating IL-22 and lower cardiovascular and cancer mortality. This aligns with UV studies showing that low-dose exposure promotes tolerogenic antigen-presenting cells and IL-10–producing regulatory T cells and dampens Th1/Th17/Th22–interferon activity, [29,30,63–65] and with observational data linking higher IL-22 to adverse cardiovascular and tumour-promoting processes. [66–71] Although there are, to our knowledge, no prior data on NHLRC3 as a UV-responsive inflammasome marker, its parallel suppression alongside IL-22 at higher Sun-BEEM exposure supports the interpretation that habitual UV exposure is associated with lower systemic inflammatory tone.

On the cardiorenal–neuroendocrine axis, FGF23 and renin are hormones involved in mineral metabolism and neurohormonal control that have been linked to cardiac hypertrophy, vascular events, and increased mortality.[72–74] Their lower levels at higher Sun-BEEM exposure, together with UVA-induced mobilisation of dermal nitric oxide and consequent vasodilation,[28,75] suggest a broader UV-driven cardiorenal–neuroendocrine pathway consistent with lower cardiovascular risk. Interestingly, previous short-term human UVB studies report that serum FGF23 rises after repeated suberythemal exposure,[76] a response generally attributed to vitamin D–driven feedback, whereas we observed lower FGF23 levels at higher habitual Sun-BEEM exposure. This contrast suggests that acute and chronic UV effects on this hormone may differ and that long-term exposure can reset the FGF23 set point despite vitamin D–induced stimulation.

For the mucosal–barrier/innate axis, higher Sun-BEEM exposure was associated with lower CD302, PIGR, LGALS4, and MMP-7, markers of innate and epithelial barrier immunity and extracellular matrix remodelling. Direct UV data on these proteins are scarce, and most prior work has described them as prognostic or tumour-related markers in haematological and epithelial cancers.[77–83] MMP-7 is the only component that has been studied in detail in the UV context, with experimental work showing increased expression in skin and other locally exposed tissues after acute or high-dose UV,[84] but UV-related changes in circulating MMP-7 have not been characterised. The inverse association between Sun-BEEM and plasma MMP-7 in our study therefore suggests spatial heterogeneity between local and systemic responses to UV and identifies a priority target for future mechanistic research.

Overall, these three axes are consistent with plausible mechanistic pathways underlying the UV–mortality associations while combining reassurance and novelty. Some components (IL-22, FGF23, renin, MMP-7) are supported by experimental and clinical data, whereas others (NHLRC3, CD302, PIGR, LGALS4) have barely been examined in the UV setting, pointing to new candidate circuits rather than simply repackaging known pathways. Across axes, the biology is largely independent of the vitamin D pathway; even FGF23, the mediator most tightly linked to vitamin D, shows the opposite pattern to short-term UVB/vitamin D trials, with lower levels at higher habitual exposure rather than the transient increases previously reported. The immunoregulatory axis has broader relevance, as systemic inflammatory tone contributes to autoimmunity, endocrine disease, and ageing, consistent with longstanding use of UV-based therapies for psoriasis and vitiligo and inverse associations between sun exposure and multiple sclerosis,[85–88] suggesting that beneficial UV effects may extend to non-cutaneous immune-mediated conditions. These mechanistic links remain hypothesis-generating, however. Finally, the contrast between short-term and long-term effects on FGF23, and between local UV-induced increases in tissue MMP-7 and lower circulating MMP-7 at higher Sun-BEEM exposure, highlights temporal and spatial heterogeneity in UV responses that future mechanistic and interventional studies should address.

### Strengths and limitations

Key strengths of this study include use of UK Biobank, a large, deeply phenotyped prospective cohort with long follow-up and near-complete linkage to national mortality and cancer registries; application of the Sun-BEEM score, a high-resolution model integrating geographic and behavioural data to estimate habitual UV exposure; and internal validation of Sun-BEEM against serum 25-hydroxyvitamin D as a biomarker of long-term sunlight exposure. A further strength is the unified modelling framework that examined cardiovascular and cancer mortality and incidence, estimated population impact fractions, and incorporated proteomic mediation analyses to explore non–vitamin D pathways. Some limitations are inherent to this observational cohort design. Residual and unmeasured confounding cannot be fully excluded, even with extensive adjustment. Selection into UK Biobank yields a healthier and more health-conscious sample than the general population, which may limit generalisability of effect sizes, although internal comparisons are likely to remain valid. UV exposure and proteomic biomarkers were measured only once at baseline, so longitudinal trajectories and temporal ordering cannot be determined. Ascertainment of some outcomes was also imperfect: non-fatal cardiovascular events were identified mainly from hospital admission and procedure codes, so milder or non-hospitalised events are likely to have been missed, and skin cancer deaths were relatively few, limiting statistical power and the precision of cause-specific estimates. This study also has method-specific limitations: Sun-BEEM was developed and calibrated entirely within UK Biobank, so external validation and adaptation are needed before similar UV exposure models can be applied in other cohorts.; In addition, The mediation analyses are based on observational data and single baseline biomarker measurements and cannot establish that the identified proteins are causal mediators; confirmation will require mechanistic and interventional studies.

## Conclusion

In this large UK cohort, higher habitual UV exposure, quantified by the Sun-BEEM index, was associated with lower all-cause, cardiovascular, and non-skin cancer mortality, alongside smaller proportional increases in melanoma and other skin cancer mortality. Population impact estimates suggest that, if these associations are causal, the reduction in cardiovascular and other cancer deaths at higher Sun-BEEM levels would outweigh the additional skin cancer deaths, indicating a net survival benefit. Proteomic mediation analyses suggest that these associations are partly mediated through three biological axes—immunoregulatory, mucosal–barrier/innate, and cardiorenal–neuroendocrine—which appear to have only a weak relation to the classical UV–vitamin D pathway and are unlikely to be reproduced by vitamin D supplementation alone. Overall, these findings challenge the simplistic view that sunlight is primarily a skin carcinogen whose benefits can be replaced by vitamin D tablets and instead support a more balanced perspective in which UV exposure contributes meaningfully, and not fully substitutable, to the prevention of cardiovascular disease and other major cancers. Future mechanistic and interventional studies are needed to define safe and effective windows of UV exposure across different populations and to test whether experimentally targeting these UV-responsive pathways can safely reduce cardiovascular and cancer risk.

## Declarations Ethics approval

UK Biobank has ethical approval from the North West Multi-centre Research Ethics Committee, and all participants provided written informed consent. This research was conducted using the UK Biobank Resource under Application Number 82526. Shortwave radiation data were provided by the Earth Observation Research Center of the Japan Aerospace Exploration Agency (JAXA).

## Patient and public involvement

Patients and the public were not involved in the design, conduct, reporting, or dissemination plans of this research.

## Data sharing

Data are available through application to the UK Biobank. Analytical code is available from the corresponding author on reasonable request.

## Transparency statement

The lead author affirms that this manuscript is an honest, accurate, and transparent account of the study being reported; that no important aspects of the study have been omitted; and that any discrepancies from the study as originally planned have been explained.

## Data Availability

The data used in this study are available from the UK Biobank (https://www.ukbiobank.ac.uk) upon reasonable request and subject to approval under the UK Biobank access procedures. This study was conducted under UK Biobank Application Number 82526.

## References

1. NCD Countdown 2030 Collaborators. NCD Countdown 2030: worldwide trends in non-communicable disease mortality and progress towards Sustainable Development Goal target 3.4. Lancet 2018;392:1072–88. doi: 10.1016/S0140-6736(18)31992-5.

2. Bray F, Laversanne M, Cao B, et al. Comparing cancer and cardiovascular disease trends in 20 middle- or high-income countries 2000–19: a pointer to national trajectories towards achieving Sustainable Development Goal target 3.4. Cancer Treat Rev 2021;100:102290. doi: 10.1016/j.ctrv.2021.102290.

3. Hoel DG, Berwick M, de Gruijl FR, et al. The risks and benefits of sun exposure. Dermatoendocrinol 2016;8:e1248325. doi: 10.1080/19381980.2016.1248325.

4. Weller RB. Sunlight: time for a rethink? J Invest Dermatol 2024;144:1724–32. doi: 10.1016/j.jid.2023.12.027.

5. NHS. How to get vitamin D from sunlight. https://www.nhs.uk/conditions/vitamins-and-minerals/vitamin-d/ (accessed 18 Dec 2025).

6. NHS. Sun safety and how to protect your skin. https://www.nhs.uk/live-well/seasonal-health/sunscreen-and-sun-safety/ (accessed 18 Dec 2025).

7. Montague M, Borland R, Sinclair C. Slip! Slop! Slap! and SunSmart, 1980–2000: skin cancer control and 20 years of population-based campaigning. Health Educ Behav 2001;28:290–305. doi: 10.1177/109019810102800304.

8. Lucas RM, Neale RE, Madronich S, et al. Are current guidelines for sun protection optimal for health? Exploring the evidence. Photochem Photobiol Sci 2018;17:1956–63. doi: 10.1039/c7pp00374a.

9. Hart PH, Norval M, Byrne SN, et al. Exposure to ultraviolet radiation in the modulation of human diseases. Annu Rev Pathol 2019;14:55–75. doi: 10.1146/annurev-pathmechdis-012418-012809.

10. Slominski RM, Raman C, Slominski AT. Photo-neuro-immuno-endocrinology: how ultraviolet radiation shapes systemic homeostasis. Proc Natl Acad Sci U S A 2024;121:e2308374121. doi: 10.1073/pnas.2308374121.

11. IARC Working Group on the Evaluation of Carcinogenic Risks to Humans. Radiation. IARC Monographs on the Evaluation of Carcinogenic Risks to Humans, Vol 100D. Lyon: International Agency for Research on Cancer; 2012.

12. Armstrong BK, Kricker A. The epidemiology of UV induced skin cancer. J Photochem Photobiol B 2001;63(1-3):8–18. doi: 10.1016/S1011-1344(01)00198-1.

13. Greinert R, de Vries E, Erdmann F, et al. European Code against Cancer 4th Edition: ultraviolet radiation and cancer. Cancer Epidemiol 2015;39(Suppl 1):S75–83. doi: 10.1016/j.canep.2014.12.014.

14. Arnold M, Singh D, Laversanne M, et al. Global burden of cutaneous melanoma in 2020 and projections to 2040. JAMA Dermatol 2022;158(5):495–503. doi: 10.1001/jamadermatol.2022.0160.

15. De Pinto G, Mignozzi S, La Vecchia C, et al. Global trends in cutaneous malignant melanoma incidence and mortality. Melanoma Res 2024;34(3):265–75. doi: 10.1097/CMR.0000000000000959.

16. Betz-Stablein B, Soyer HP. Overdiagnosis in melanoma screening: is it a real problem? Dermatol Pract Concept 2023;13(4):e2023247. doi: 10.5826/dpc.1304a247.

17. Adamson AS, Suarez EA, Welch HG. Estimating overdiagnosis of melanoma using trends among Black and White patients in the US. JAMA Dermatol 2022;158(4):426–31. doi: 10.1001/jamadermatol.2022.0139.

18. Whiteman DC. Testing the divergent pathway hypothesis for melanoma: recent findings and future challenges. Expert Rev Anticancer Ther 2010;10(5):615–8. doi: 10.1586/era.10.42.

19. Whiteman DC, Watt P, Purdie DM, et al. Melanocytic nevi, solar keratoses, and divergent pathways to cutaneous melanoma. J Natl Cancer Inst 2003;95(11):806–12. doi: 10.1093/jnci/95.11.806.

20. Grant WB. An estimate of premature cancer mortality in the United States due to inadequate doses of solar ultraviolet-B radiation. Cancer 2002;94(6):1867–75. doi: 10.1002/cncr.10427.

21. Freedman DM, Dosemeci M, McGlynn K. Residential and occupational exposure to sunlight and mortality from non-Hodgkin’s lymphoma: composite (threefold) case-control study. BMJ 1997;314:1451–5.

22. Olsen CM, Pandeya N, Law MH, et al. Does polygenic risk influence associations between sun exposure and melanoma? A prospective cohort analysis. Br J Dermatol 2020;183:303–10. doi: 10.1111/bjd.18703.

23. Chowdhury R, Kunutsor S, Vitezova A, et al. Vitamin D and risk of cause specific death: systematic review and meta-analysis of observational cohort and randomised intervention studies. BMJ 2014;348:g1903. doi:10.1136/bmj.g1903.

24. Manson JE, Cook NR, Lee IM, et al. Vitamin D supplements and prevention of cancer and cardiovascular disease. N Engl J Med 2019;380:33–44. doi: 10.1056/NEJMoa1809944.

25. Zhang Y, Fang F, Tang J, et al. Association between vitamin D supplementation and mortality: systematic review and meta-analysis. BMJ 2019;366:l4673. doi: 10.1136/bmj.l4673.

26. Afzal S, Brøndum-Jacobsen P, Bojesen SE, et al. Genetically low vitamin D concentrations and increased mortality: mendelian randomisation analysis in three large cohorts. BMJ 2014;349:g6330. doi: 10.1136/bmj.g6330.

27. Opländer C, Volkmar CM, Paunel-Görgülü A, et al. Whole-body UVA irradiation lowers systemic blood pressure by release of nitric oxide from intracutaneous photolabile nitric oxide derivatives. Circ Res 2009;105:1031–40. doi: 10.1161/CIRCRESAHA.109.207019.

28. Liu D, Fernandez BO, Hamilton A, et al. UVA irradiation of human skin vasodilates arterial vasculature and lowers blood pressure independently of nitric oxide synthase. J Invest Dermatol 2014;134:1839–46. doi: 10.1038/jid.2014.27.

29. Hart PH, Gorman S, Finlay-Jones JJ. Modulation of the immune system by UV radiation: more than just the effects of vitamin D? Nat Rev Immunol 2011;11:584–96. doi:10.1038/nri3045.

30. Bernard JJ, Krutmann J, Gallo RL. Photoimmunology: how ultraviolet radiation affects the immune system. Nat Rev Immunol 2019;19:688–701. doi:10.1038/s41577-019-0185-9.

31. Vieyra-Garcia PA, Wolf P. A deep dive into UV-based phototherapy: mechanisms of action and emerging molecular targets in inflammation and cancer. Pharmacol Ther 2021;222:107784. doi: 10.1016/j.pharmthera.2020.107784.

32. Lindqvist PG, Epstein E, Landin-Olsson M, et al. Avoidance of sun exposure as a risk factor for major causes of death: a competing risk analysis of the Melanoma in Southern Sweden cohort. J Intern Med 2016;280:375–87. doi:10.1111/joim.12496.

33. Stevenson AC, Clemens T, Pairo-Castineira E, et al. Higher ultraviolet light exposure is associated with lower mortality: an analysis of data from the UK Biobank cohort study. Health Place 2024;89:103328. doi:10.1016/j.healthplace.2024.103328.

34. Nazeeh N, Orlich MJ, Segovia-Siapco G, et al. The association between time spent outdoors during daylight and mortality among participants of the Adventist Health Study 2 Cohort. Environ Epidemiol 2025;9:e401. doi:10.1097/EE9.0000000000000401.

35. Lindqvist PG, Epstein E, Landin-Olsson M. Sun Exposure - Hazards and Benefits. Anticancer Res 2022;42:1671–7. doi:10.21873/anticanres.15644.

36. Keil AP, Edwards JK, Richardson DB, et al. The parametric g-formula for time-to-event data: intuition and a worked example. Int J Epidemiol 2014;43:1760–71. doi:10.1093/ije/dyu187.

37. Mansournia MA, Altman DG. Population attributable fraction. BMJ 2018;360:k757. doi:10.1136/bmj.k757.

38. Sudlow C, Gallacher J, Allen N, et al. UK Biobank: an open access resource for identifying the causes of a wide range of complex diseases of middle and old age. PLoS Med 2015;12:e1001779. doi:10.1371/journal.pmed.1001779.

39. Bycroft C, Freeman C, Petkova D, et al. The UK Biobank resource with deep phenotyping and genomic data. Nature 2018;562:203–209. doi:10.1038/s41586-018-0579-z.

40. Scragg R, Camargo CA Jr. Frequency of leisure-time physical activity and serum 25-hydroxyvitamin D levels in the US population: results from the Third National Health and Nutrition Examination Survey. Am J Epidemiol 2008;168:577–86. doi:10.1093/aje/kwn163.

41. Boniol M, Autier P, Boyle P, et al. Cutaneous melanoma attributable to sunbed use: systematic review and meta-analysis. BMJ 2012;345:e4757. doi:10.1136/bmj.e4757.

42. Autier P, Boniol M, Doré JF. Sunscreen use and increased duration of intentional sun exposure: still a burning issue. Int J Cancer 2007;121:1–5. doi:10.1002/ijc.22745.

43. Lindqvist PG, Epstein E, Landin-Olsson M, et al. Avoidance of sun exposure is a risk factor for all-cause mortality: results from the Melanoma in Southern Sweden cohort. J Intern Med 2014;276:77–86. doi:10.1111/joim.12251.

44. He W, Zhu F, Ma X, et al. Actinic skin damage and mortality: the First National Health and Nutrition Examination Survey Epidemiologic Follow-Up Study. PLoS One 2011;6:e19907. doi:10.1371/journal.pone.0019907.

45. Christoffersen M, Frikke-Schmidt R, Schnohr P, et al. Visible age-related signs and risk of ischemic heart disease, myocardial infarction, and death in the general population. Circulation 2014;129:990–8. doi:10.1161/CIRCULATIONAHA.113.001696.

46. Esquirol Y, Ferrieres J, Marquie JC, et al. Forehead wrinkles and risk of all-cause and cardiovascular mortality over 20-year follow-up in a working population: the VISAT study. Eur Heart J 2018;39(suppl 1):ehy565.P1548. doi:10.1093/eurheartj/ehy565.P1548.

47. Newton-Bishop J, Chang YM, Elliott F, et al. Melanocytic nevi, nevus genes, and melanoma risk in a large case-control study in the United Kingdom. Cancer Epidemiol Biomarkers Prev 2010;19:2043–54. doi:10.1158/1055-9965.EPI-10-0233.

48. Cancer Research UK. Non-melanoma skin cancer mortality statistics. Available at: https://www.cancerresearchuk.org/health-professional/cancer-statistics/statistics-by-cancer-type/non-melanoma-skin-cancer Accessed 18 Dec 2025.

49. Cancer Research UK. Risks and causes of melanoma skin cancer. Available at: https://www.cancerresearchuk.org/about-cancer/melanoma/risks-causes Accessed 18 Dec 2025.

50. Parkin DM, Mesher D, Sasieni P. Cancers attributable to exposure to solar (ultraviolet) radiation in the UK in 2010. Br J Cancer 2011;105 Suppl 2:S66–9. doi:10.1038/bjc.2011.486.

51. Welch HG, Mazer BL, Adamson AS. The rapid rise in cutaneous melanoma diagnoses. N Engl J Med 2021;384:72–9. doi:10.1056/NEJMsb2019760.

52. Oke JL, Nicholson BD, Lay-Flurrie S, et al. Mapping cancer incidence and mortality in the UK: 1980-2013. Sci Rep 2018;8:14663. doi:10.1038/s41598-018-32900-6.

53. Nielsen JB, Kristiansen IS, Thapa S. Increasing melanoma incidence with unchanged mortality: more sunshine, better treatment, increased diagnostic activity, overdiagnosis or lowered diagnostic threshold? Br J Dermatol 2024;191(3):365–374. doi:10.1093/bjd/ljae175.

54. Weller RB, Gu J. Ultraviolet radiation is not the major cause of melanoma mortality in the UK and sun exposure advice should be revised. Br J Dermatol 2025;192:548–50. doi:10.1093/bjd/ljae426.

55. British Heart Foundation. Heart & circulatory disease statistics 2024. Available at: https://www.bhf.org.uk/what-we-do/our-research/heart-statistics/heart-statistics-publications/cardiovascular-disease-statistics-2024 Accessed 18 Dec 2025.

56. National Institute for Health and Care Excellence. CVD risk assessment and management: burden of cardiovascular disease. https://cks.nice.org.uk/topics/cvd-risk-assessment-management/background-information/burden-of-cvd/. Accessed 18 Dec 2025.

57. Cancer Research UK. Melanoma skin cancer mortality statistics. https://www.cancerresearchuk.org/health-professional/cancer-statistics/statistics-by-cancer-type/melanoma-skin-cancer/mortality. Accessed 18 Dec 2025.

58. Luengo-Fernandez R, Gray AM, Bull L, et al. Quality of life after TIA and stroke: ten-year results of the Oxford Vascular Study. Neurology 2013;81(18):1588–95. doi:10.1212/WNL.0b013e3182a9f45f.

59. Pettersen KI, Reikvam A, Rollag A, et al. Health-related quality of life after myocardial infarction is associated with left ventricular ejection fraction. BMC Cardiovasc Disord 2008;8:28. doi:10.1186/1471-2261-8-28.

60. National Institute for Health and Care Excellence. Pembrolizumab for advanced melanoma not previously treated with ipilimumab. NICE technology appraisal guidance TA357. London: NICE; 2015. Accessed 18 Dec 2025

61. Tennvall GR, Norlin JM, Malmberg I, et al. Health-related quality of life in patients with actinic keratosis. Acta Derm Venereol 2015;95:193–9. doi:10.2340/00015555-1919.

62. Philipp-Dormston WG, Müller K, Novak N, et al. Patient-reported health outcomes in patients with non-melanoma skin cancer and actinic keratosis: a large observational study. J Eur Acad Dermatol Venereol 2018;32:1110–6. doi:10.1111/jdv.14830.

63. Schwarz T. Ultraviolet radiation-induced tolerance. Allergy 1999;54:1252–61. doi:10.1034/j.1398-9995.1999.00105.x.

64. Rácz E, Prens EP, Kurek D, et al. Effective treatment of psoriasis with narrow-band UVB phototherapy is linked to suppression of the IFN and Th17 pathways. J Invest Dermatol 2011;131:1547–58. doi:10.1038/jid.2011.53.

65. Furio L, Berthier-Vergnes O, Ducarre B, et al. UVA radiation impairs phenotypic and functional maturation of human dermal dendritic cells. J Invest Dermatol 2005;125(5):1032–8. doi:10.1111/j.0022-202X.2005.23904.x.

66. Lewis JM, Monico PF, Mirza FN, et al. Chronic UV radiation-induced RORγt+ IL-22-producing lymphoid cells are associated with mutant KC clonal expansion. Proc Natl Acad Sci U S A 2021;118:e2016963118. doi:10.1073/pnas.2016963118.

67. Luo JW, Hu Y, Liu J, et al. Interleukin-22: a potential therapeutic target in atherosclerosis. Mol Med 2021;27(1):88. doi:10.1186/s10020-021-00353-9.

68. Ye J, Ji Q, Liu J, et al. Interleukin 22 promotes blood pressure elevation and endothelial dysfunction in angiotensin II-treated mice. J Am Heart Assoc 2017;6(10):e005875. doi:10.1161/JAHA.117.005875.

69. Deng H, Li H, Liu Z, Shen N, Dong N, Deng C, et al. Pro-osteogenic role of interleukin-22 in calcific aortic valve disease. Atherosclerosis 2024;388:117424. doi:10.1016/j.atherosclerosis.2023.117424

70. Lim C, Savan R. The role of the IL-22/IL-22R1 axis in cancer. Cytokine Growth Factor Rev 2014;25(3):257–71. doi:10.1016/j.cytogfr.2014.04.005.

71. Voigt C, May P, Gottschlich A, et al. Cancer cells induce interleukin-22 production from memory CD4+ T cells via interleukin-1 to promote tumor growth. Proc Natl Acad Sci U S A 2017;114:12994–9. doi:10.1073/pnas.1705165114.

72. Qin Z, Liu X, Song M, Zhou Q, Yu J, Zhou B, et al. Fibroblast growth factor 23 as a predictor of cardiovascular and all-cause mortality in prospective studies. Atherosclerosis 2017;261:1–11. doi:10.1016/j.atherosclerosis.2017.03.042.

73. Verma S, Gupta M, Holmes DT, et al. Plasma renin activity predicts cardiovascular mortality in the Heart Outcomes Prevention Evaluation (HOPE) study. Eur Heart J 2011;32:2135–42. doi:10.1093/eurheartj/ehr066.

74. Bhandari SK, Batech M, Shi J, et al. Plasma renin activity and risk of cardiovascular and mortality outcomes among individuals with elevated and nonelevated blood pressure. Kidney Res Clin Pract 2016;35:219–28. doi:10.1016/j.krcp.2016.07.004.

75. Halliday GM, Damian DL. An unexpected role: UVA-induced release of nitric oxide from skin may have unexpected health benefits. J Invest Dermatol 2014;134:1797–9. doi:10.1038/jid.2014.170.

76. Lee JH, Kim KM, Shin DY, et al. Ultraviolet B activated 1,25(OH)2D affects the level of fibroblast growth factor-23 in human. Endocr J 2013;60:81–6. doi:10.1507/endocrj.EJ12-0199.

77. Lo T-H, Silveira PA, Fromm PD, et al. Characterization of the expression and function of the C-type lectin receptor CD302 in mice and humans reveals a role in dendritic cell migration. J Immunol 2016;197:885–98. doi:10.4049/jimmunol.1600259.

78. Kato M, Khan S, d’Aniello E, et al. The novel endocytic and phagocytic C-type lectin receptor DCL-1/CD302 on macrophages is colocalized with F-actin, suggesting a role in cell adhesion and migration. J Immunol 2007;179:6052–63. doi:10.4049/jimmunol.179.9.6052.

79. Alaterre E, Raimbault S, Goldschmidt H, et al. CD24, CD27, CD36 and CD302 gene expression for outcome prediction in patients with multiple myeloma. Oncotarget 2017;8:98931–44. doi:10.18632/oncotarget.22131.

80. Jaffar Z, Ferrini ME, Herritt LA, et al. Cutting edge: lung mucosal Th17-mediated responses induce polymeric Ig receptor expression by the airway epithelium and elevate secretory IgA levels. J Immunol 2009;182:4507–11. doi:10.4049/jimmunol.0900237.

81. Fristedt R, Gaber A, Hedner C, et al. Expression and prognostic significance of the polymeric immunoglobulin receptor in esophageal and gastric adenocarcinoma. J Transl Med 2014;12:83. doi:10.1186/1479-5876-12-83.

82. Cao AT, Yao S, Gong B, et al. Th17 cells upregulate polymeric Ig receptor and intestinal IgA and contribute to intestinal homeostasis. J Immunol 2012;189:4666–73. doi:10.4049/jimmunol.1200955.

83. Moreno-Ajona D, Irimia P, Rodríguez JA, et al. Elevated circulating metalloproteinase 7 predicts recurrent cardiovascular events in patients with carotid stenosis: a prospective cohort study. BMC Cardiovasc Disord 2020;20:93. doi:10.1186/s12872-020-01387-3.

84. Quan T, Qin Z, Xia W, et al. Matrix-degrading metalloproteinases in photoaging. J Investig Dermatol Symp Proc 2009;14:20–4. doi:10.1038/jidsymp.2009.8.

85. Elmets CA, Lim HW, Stoff B, et al. Joint American Academy of Dermatology–National Psoriasis Foundation guidelines of care for the management and treatment of psoriasis with phototherapy. J Am Acad Dermatol 2019;81:775–804. doi:10.1016/j.jaad.2019.04.042.

86. Goulden V, Ling TC, Babakinejad P, et al. British Association of Dermatologists and British Photodermatology Group guidelines for narrowband ultraviolet B phototherapy 2022. Br J Dermatol 2022;187:295–308. doi:10.1111/bjd.21669.

87. van der Mei IAF, Ponsonby A-L, Dwyer T, et al. Past exposure to sun, skin phenotype, and risk of multiple sclerosis: case-control study. BMJ 2003;327:316. doi:10.1136/bmj.327.7410.316.

88. Bäärnhielm M, Hedström AK, Kockum I, et al. Sunlight is associated with decreased multiple sclerosis risk: no interaction with human leukocyte antigen-DRB1*15. Eur J Neurol 2012;19:955–62. doi:10.1111/j.1468-1331.2011.03650.x.

