## Supplementary Methods, Tables and Figures for "Risk–benefit balance of habitual ultraviolet exposure for cardiovascular, cancer, and skin cancer mortality: UK Biobank cohort study"

**1. Study population and analytic cohorts**

UK Biobank is a large population-based prospective cohort of 502 492 adults aged 40-70 years, recruited from 22 assessment centres across England, Scotland, and Wales between 2006 and 2010. All participants completed touchscreen questionnaires on sociodemographic factors, lifestyle, and medical history, underwent physical measurements, and provided biological samples, with long term follow-up through linkage to National Health Service (NHS) electronic health records [1].

For the present analyses, we restricted to participants of genetically confirmed European ancestry to reduce heterogeneity in UV responses related to skin pigmentation. After excluding individuals without complete data on habitual UV exposure or follow-up, the main analytic cohort comprised 419 007 participants with information on the Sun-Behavioural and Environmental Exposure Model (Sun-BEEM) score and mortality outcomes.

A subset of UK Biobank participants had plasma proteomic profiling under the UK Biobank Pharma Proteomics Project, using Olink’s Explore 1536 proximity extension assay (PEA) with next-generation sequencing readout[2]. After quality control and restriction to those with Sun-BEEM and follow-up data, 44 712 participants formed the proteomic subcohort used for mediation analyses.

**2. Assessment of habitual UV exposure: the Sun-BEEM score**

**2.1 Conceptual rationale and design**

The design of Sun-BEEM was guided by three considerations. First, we aimed to incorporate both environmental potential for UV exposure (ambient shortwave radiation) and behavioural components (time spent outdoors, use of artificial UV sources, and sun/UV protection), because experimental and epidemiological work indicates that systemic effects of UV depend on the combination of dose delivered at ground level and individual behaviour[3]. These components were selected because they are both readily available in UK Biobank and have each been examined previously as indicators of UV exposure.

Second, we prioritised robust, interpretable categories over fine-grained but noisy quantitative measures. Self-reported time outdoors, solarium/sunlamp use and sun/UV protection are prone to recall and reporting error and are not naturally measured on a precise continuous scale; solarium/sunlamp use is relatively rare in UK Biobank. Using simple binary codings within each domain therefore reduces model complexity and avoids unstable estimates in small extreme groups[4].

Third, we constructed an unweighted composite score by summing the four binary components. There was no strong prior evidence on the relative contributions of environmental versus behavioural components to long term systemic effects of UV exposure, and deriving weights from the same dataset would risk overfitting and circularity. Unweighted composite lifestyle and exposome scores—defined as the simple sum of individual low- or high-risk components—are widely used in large-scale cohort studies and have been shown to predict cardiovascular, cancer and other chronic disease outcomes [5] [6].We therefore chose an analogous approach for habitual UV exposure.

In exploratory analyses, we also examined data-driven UV exposure patterns. We fitted latent class analysis (LCA) models to the four binary Sun-BEEM components, allowing two to six classes. We evaluated solutions using information criteria, class sizes and the substantive interpretability of latent phenotypes [7] [8] Across this range, several classes were very small and profile overlap was substantial, so no solution yielded clearly interpretable UV exposure classes; We also applied unsupervised clustering (k-means and Gaussian mixture models) to the underlying continuous/ranking measures (ambient shortwave radiation, average time outdoors, frequency of solarium/sunlamp use, and frequency of sun/UV protection). For all reasonable choices of the number of clusters (k = 2–6), average silhouette values were close to zero and cluster profiles were highly overlapping, indicating that the data did not support stable, distinct groupings[9]. Because these data-driven approaches failed to produce well-separated or interpretable phenotypic clusters, we did not pursue them further (details available on request), and retained the prespecified, unweighted Sun-BEEM scorer.

**2.2 Components and construction**

**2.2.1 Ambient shortwave radiation**

Annual downward shortwave radiation (SWR, kJ/m²) spaning 300 nm–4 µm was estimated for each participant’s residential address in 2008 using satellite-based MODIS Terra/Aqua surface radiation products (MCD18 series), which provide daily combined downward shortwave radiation fields on a global grid[10]. Residential coordinates supplied by UK Biobank (1 km × 1 km Ordnance Survey grid) were converted to latitude–longitude and matched to the nearest grid cell. We calculated annual mean SWR for each grid cell by averaging monthly means for 2008, chosen as a representative mid-recruitment year (2006–2010).

Participants living in areas with SWR below an a priori cut-off approximating the median at a centrally located assessment centre (Nottingham, 9516 kJ/m²) were coded as low ambient solar radiation (component score = 0); those at or above the cut-off were coded as high ambient solar radiation (score = 1). Median splits of continuous environmental exposures are commonly used in large-scale composite lifestyle and exposome scores when no established biological risk threshold exists and when interpretability and comparability across settings are priorities [5] [6].

**2.2.2 Habitual time spent outdoors**

Participants reported typical hours spent outdoors on a summer and a winter day; these were averaged to yield a year-round measure of habitual time spent outdoors (hours/day). We dichotomised this variable at ≥4 hours/day versus <4 hours/day.

The 4-hour threshold was chosen a priori to represent a “high exposure” pattern in high-latitude countries, while retaining adequate numbers of participants in each group. Meta-analytic and survey data from northern European and other temperate populations indicate that most adults spend around 1–2 hours/day outdoors, with right-skewed distributions and only a minority reporting ≥3–4 hours/day across the year or during the summer season[11] [12] [13]. In these studies, ~4 hours/day typically lies in the upper tail of the distribution and is more characteristic of people with outdoor-intensive occupations, frequent recreational outdoor activities, or sun-seeking holidays than of the general working-age population.

On this basis, we used 4 hours/day as a pragmatic cut-off to distinguish participants whose habitual outdoor time is clearly above the norm (and therefore likely to accumulate substantially higher personal UV exposure) from the majority of the cohort. Participants with <4 hours/day were coded as low behavioural exposure (component score = 0) and those with ≥4 hours/day as high (score = 1), yielding a simple, interpretable contrast between “typical” and “high” outdoor exposure within the UK Biobank setting.

**2.2.3 Solarium / sunlamp use**

Self-reported frequency of solarium/sunlamp use was dichotomised as ≥1 time/year versus <1 time/year or never, to capture occasional or regular use of high-intensity artificial UV sources. “Do not know” and “prefer not to answer” were set to missing.

Epidemiological studies of indoor tanning and skin cancer risk commonly use an initial contrast of ever versus never use of sunbeds or sunlamps as the main exposure comparison [14] [15] [16]. Therefore, dichotomising solarium/sunlamp use as ≥1 time per year versus <1 time per year or never follows established practice and separates a behaviourally distinct group with non-negligible additional artificial UV exposure from non-users. Notably, solarium use was generally infrequent with most users reporting 1–6 sessions per year and likely reflects broader sun-seeking behaviour.

**2.2.4 Sun/UV protection**

Participants reported how often they used sun/UV protection (e.g. sunscreen, protective clothing) when outdoors in summer. We treated this variable primarily as a marker of “sun-reactive” behaviour—combining some degree of sun sensitivity with intentional management of outdoor exposure—rather than as a simple inverse measure of dose. Participants who reported using protection “sometimes”, “most of the time” or “always” were coded as 1, indicating that they anticipate enough sun exposure to consider protection; those who answered “never/rarely” or “do not go out in sunshine” were coded as 0.

Epidemiological and behavioural studies suggest that sunscreen use is often highest in individuals who seek or tolerate more sun exposure, including sunbathers, “sun worshippers”, and those taking sunny holidays, and that sunscreen use correlates with risk behaviours such as exposing the upper body or deliberately staying longer in the sun. In several observational studies and randomised trials, sunscreen users—particularly those using higher SPF products—spent longer in the sun than non-users or low-SPF users when exposure was intentional. [17] [18] Behavioural surveys and reviews further note that positive attitudes to suntanning, high sun exposure, and sunscreen use frequently cluster together[18] [19]. In this context, our coding is intended to distinguish participants who have essentially minimal intentional sun exposure (“never/rarely” or “do not go out in sunshine”) from those with at least some planned time outdoors who report adopting protective measures, while recognising that protection may partially mitigate but does not eliminate UV dose.

**2.2.5 Sun-BEEM score and categories**

Each of the four components (ambient SWR, habitual time outdoors, solarium/sunlamp use, sun/UV protection) was coded as a binary indicator (0/1), and the Sun-BEEM score was calculated as their unweighted sum (range 0–4), with higher values indicating higher cumulative habitual UV exposure. Participants with missing data on any of the four Sun-BEEM components were excluded from analyses that used the Sun-BEEM score, to avoid ambiguity about overall exposure classification.

For the main analyses, Sun-BEEM was grouped into three categories, consistent with the main text: low UV (score 0–1), medium UV (score 2), and high UV (score 3–4). This grouping preserves the ordinal structure of the score while avoiding sparse categories at extreme values, analogous to unweighted composite lifestyle scores that are commonly categorised into low, intermediate and high groups to improve stability and interpretability of risk estimates.

**2.3 Empirical validation and performance**

To evaluate construct validity, we examined cross-sectional associations between Sun-BEEM and two markers expected to reflect long-term UV exposure: serum 25-hydroxyvitamin D [25(OH)D], measured at baseline, and self-reported number of childhood sunburns. UV-B exposure is the principal determinant of endogenous vitamin D synthesis[20], whereas childhood sunburns reflect high-intensity intermittent UV exposure, which is epidemiologically linked to adult skin cancer risk[21].

We compared mean 25(OH)D and mean childhood sunburn counts across Sun-BEEM categories using linear models adjusted for age, sex, BMI, season and assessment centre. As a further check, we examined the individual binary components of Sun-BEEM. For each component (high vs low SWR, ≥4 vs <4 hours/day outdoors, solarium/sunlamp use vs no use, high vs low sun/UV protection), we compared mean 25(OH)D and mean childhood sunburn counts across exposure strata using t tests or linear regression. These results are summarised in Supplementary Tables S5 and S6.

In addition, we assessed the sensitivity of associations to alternative codings of the underlying components (for example, treating ambient SWR and outdoor time as continuous variables, or modelling the 0–4 Sun-BEEM score without grouping). These alternative specifications produced very similar patterns and magnitudes of association with both validation markers and the main mortality and incident outcomes (data available on request).

Taken together, higher Sun-BEEM categories and higher values of the individual components showed the expected positive gradients in 25(OH)D and childhood sunburn, and alternative codings yielded consistent results. These findings support the use of Sun-BEEM as a simple, interpretable summary of habitual UV exposure in this cohort.

**3. Covariates and handling of missing data**

**3.1 Covariate definitions**

Based on prior literature and causal reasoning, we identified a set of a priori covariates that could confound associations between UV exposure and health outcomes. These included sociodemographic, anthropometric and lifestyle factors. Sociodemographic factors were age at baseline, grouped into approximately equal-sized bands derived from the age distribution in the analytic cohort (37–47, 48–54, 55–59, 60–63, 64–73 years); sex; Townsend deprivation index (quintiles); and highest educational attainment (less than secondary, secondary school, university or above)[22] . Anthropometry was represented by BMI, derived from measured height and weight, categorised as underweight (<18.5 kg/m²), healthy weight (18.5–<25), overweight (25–<30), and obese (≥30) using standard clinical thresholds, in line with World Health Organization and UK clinical definitions of adult BMI categories.

Lifestyle behaviours included smoking status (current, previous, never)[23]; alcohol intake (derived in UK units per day from self-reported drinking frequency and typical quantity, and dichotomised as within (≤14 units per week) versus above (>14 units per week) the UK Chief Medical Officers’ recommended limits)[24]; physical activity (total weekly metabolic equivalent [MET] minutes, grouped as low <600, moderate 600–2999, high ≥3000) [25] [26]; and a composite sleep score capturing chronotype, sleep duration, insomnia, snoring, and daytime sleepiness, derived from five binary indicators and classified as healthy (4–5 favourable dimensions), intermediate (2–3), or poor (0–1), as in prior UK Biobank “healthy sleep” indices[27].Full operational definitions and UK Biobank field IDs are provided in Supplementary Table S3.

**3.2 Missing data and multiple imputation**

Participants with missing data on the Sun-BEEM exposure or on outcome variables were excluded from the analyses and were not imputed. For continuous variables used to define categories (body mass index and Townsend deprivation score), occasional missing values were replaced with the sample mean prior to categorisation. For the five binary components of the composite sleep score, missing values were imputed by random draws from the observed distribution of each component (proportional imputation). For the remaining sociodemographic and lifestyle covariates, missing categories were imputed under a missing-at-random assumption using multivariate imputation by chained equations (MICE; random forest models, five imputed datasets), with primary analyses based on one completed dataset and complete-case analyses used as sensitivity checks[28] [29].

**4. Outcome definitions and coding**

**4.1 Mortality outcomes**

Mortality was ascertained through linkage to national death registries (NHS Digital for England and Wales; NHS Central Register for Scotland), using ICD-10 codes.[1] We examined all-cause mortality (any death recorded after baseline), cardiovascular mortality (narrow definition: primary cause codes I21–I25 for ischaemic heart disease or I60–I64 for stroke)[30], cancer mortality (primary cause C00–C97 or D37–D48)[31], and skin cancer mortality(melanoma mortality was defined as deaths with primary cause C43, and non-melanoma skin cancer mortality as deaths with primary cause C44). Non-skin cancer mortality was defined as cancer mortality excluding C43 and C44.

Follow-up time for mortality was calculated from baseline assessment date to date of death or censoring at the latest registry linkage (31 March 2025). Participants known to be alive at this date were censored; those with incomplete linkage were excluded from cause-specific analyses. Detailed ICD-10 code lists and UK Biobank field IDs used for each mortality and incident outcome definition are provided in Supplementary Table S4.

**4.2 Incident cardiovascular and cancer outcomes**

Incident cancer outcomes were ascertained through linkage to national cancer registries, with diagnoses coded using ICD-10.[1] Any incident cancer was defined as the first registration with a primary malignant neoplasm (C00–C97) or neoplasm of uncertain or unknown behaviour (D37–D48). Incident melanoma was defined as the first registration with cutaneous melanoma (C43), incident non-melanoma skin cancer as the first registration with keratinocyte carcinoma (C44), and incident non-skin cancer as the first registration with cancer excluding C43 and C44.

Incident cardiovascular disease (CVD) events were identified from linked hospital inpatient records (Hospital Episode Statistics for England, Welsh hospital datasets, and the Scottish Morbidity Record), using the same ICD-10 code ranges as for cardiovascular mortality.[1] Incident major CVD was defined as the first hospital admission with a primary or secondary diagnosis of ischaemic heart disease (I21–I25) or stroke (I60–I64).

Participants with evidence of the corresponding disease prior to the baseline assessment were excluded from incident analyses. Specifically, for cancer outcomes, individuals with any cancer registration before baseline were excluded from analyses of any incident cancer and site-specific incident cancers. For CVD outcomes, individuals with any hospital inpatient record with relevant ICD-10 codes (I21–I25 or I60–I64) recorded before baseline were excluded from the incident major CVD analysis. Baseline was defined as the date of attendance at the UK Biobank assessment centre.

For all incident outcomes, follow-up time was calculated from baseline assessment to the date of first qualifying event, death, loss to follow-up, or the end of linkage for the relevant hospital or cancer registry dataset, whichever occurred first. Participants without an event were censored at the last available linkage date. Detailed code lists, data sources, and censoring dates for each incident endpoint are provided in Supplementary Table S4.

**5. Plasma proteomics**

Baseline EDTA plasma samples were collected at assessment centres and processed under standardised UK Biobank protocols[32]. Proteomic profiling was performed using the Olink® Explore 1536 platform (Olink Proteomics AB, Sweden), which measures approximately 1500 proteins across four panels using proximity extension assays with next-generation sequencing readout[33].

Protein abundances were expressed on the Normalized Protein Expression (NPX, log₂) scale, after central normalisation and batch correction within UK Biobank’s proteomics pipeline. We restricted analyses to proteins with acceptable quality control and detection in at least 80% of samples. Values below limits of detection were handled according to platform-recommended procedures (for example, limit-of-detection–based replacement or censoring rules)[34].

The final proteomic dataset for mediation analyses comprised 44 712 participants with complete Sun-BEEM, covariate, mortality, and proteomic data, and 2 494 proteins that passed these quality filters.

**6. Statistical analysis**

All analyses were conducted using R version 4.4.0 (R Foundation for Statistical Computing, Vienna, Austria)[35].

**6.1 Descriptive analyses**

Baseline characteristics were summarised across Sun-BEEM categories (low, medium, high) using means (standard deviations) for continuous variables and counts (percentages) for categorical variables. Differences across exposure groups were assessed using one-way ANOVA for continuous variables and χ² tests for categorical variables; the full baseline table is presented in the main manuscript. Associations of Sun-BEEM and its binary components with serum 25(OH)D and childhood sunburn are summarised in Supplementary Tables S5 and S6, as described in Section 2.3.

**6.2 Main Cox models for mortality and incidence**

Associations between Sun-BEEM categories and time-to-event outcomes were estimated using Cox proportional hazards models[36] [37], with time since baseline as the underlying timescale. For each outcome, we fitted a crude model including Sun-BEEM only and a multivariable model including Sun-BEEM and all a priori covariates (age group, sex, BMI category, Townsend deprivation quintile, educational attainment, smoking status, alcohol intake, physical activity category, sleep score). Sun-BEEM low served as the reference category; hazard ratios (HRs) and 95% confidence intervals (CIs) were reported for medium and high categories compared with low. Supplementary Figure S1 (incident outcomes) and Supplementary Figure S2 (mortality outcomes), illustrating that the inverse associations between higher Sun-BEEM and internal causes of death were not driven by adjustment for any single covariate block.

**6.3 Functional form and proportional hazards assumption**

**6.3.1 Restricted cubic spline analyses**

To explore potential non-linear dose–response relationships between continuous UV-related variables and mortality, we fitted Cox models with solarium use frequency (times/year), ambient SWR (kJ/m²) and average outdoor time (hours/day), modelled using restricted cubic splines with four knots, implemented via the rms package[38]. For each outcome–exposure pair, we compared a spline model against a simpler linear model for the exposure using likelihood ratio tests or partial Wald tests via anova.cph. Evidence of non-linearity was judged based on the joint significance of the non-linear spline terms. Formal test statistics and detailed curves are not tabulated but were examined to ensure that the chosen categorical Sun-BEEM specification captured the main patterns; additional information is available on request.

**6.3.2 Proportional hazards assumption**

We examined the proportional hazards assumption using Schoenfeld residuals[39].For each main Cox model, we applied cox.zph to obtain global and covariate-specific tests and visual plots of scaled Schoenfeld residuals against time. Where global tests indicated no strong violation, we retained standard Cox models. For models with borderline evidence of non-proportionality, residual plots showed only minor deviations and did not materially alter Sun-BEEM HRs. Because results were consistent with the main analyses, we did not provide a separate supplementary table of Schoenfeld test statistics.

**6.4 Population impact fractions via parametric g-computation**

To quantify the potential population-level impact of altering habitual UV exposure, we used parametric g-computation (the parametric g-formula) based on the multivariable Cox models [40] [41] . For each cause-specific mortality outcome (k), we considered the observed Sun-BEEM distribution, a counterfactual scenario in which all participants were assigned Sun-BEEM low (score 0–1; Scenario A, low UV), and a counterfactual scenario in which all participants were assigned Sun-BEEM high (score 3–4; Scenario B, high UV).

Using the fitted Cox models and observed covariates, we predicted the expected number of deaths under each scenario over the observed follow-up:$D_{k}^{\text{obs}}$, the expected number of deaths under the observed Sun-BEEM distribution; $D_{k}^{\text{low}}$ , the expected number of deaths if all participants were assigned Sun-BEEM low; and $D_{k}^{\text{high}}$, the expected number of deaths if all participants were assigned Sun-BEEM high. Population impact fractions (PIFs, equivalently attributable fractions) were defined as:

$$AF_{k}^{\left( A \right)} = \frac{D_{k}^{\text{obs}} - D_{k}^{\text{low}}}{D_{k}^{\text{obs}}}$$

for Scenario A (all low UV); and

$$AF_{k}^{\left( B \right)}=\frac{D_{k}^{\text{obs}}-D_{k}^{\text{high}}}{D_{k}^{\text{obs}}}$$

for Scenario B (all high UV). Positive values indicate fewer deaths under the counterfactual (a protective shift), whereas negative values indicate more deaths (a harmful shift). We did not assume a priori which Sun-BEEM category conferred lower risk for any given outcome.

Absolute impacts were expressed as attributable deaths and excess deaths: (for protective scenarios: $AD_{k}=D_{k}^{\text{obs}}-D_{k}^{\text{cf}}$ ) and excess deaths (for harmful scenarios: $ED_{k}=D_{k}^{\text{cf}}-D_{k}^{\text{obs}}$ ) , where $D_{k}^{\text{cf}}$ denotes $D_{k}^{\text{low}}$ or $D_{k}^{\text{high}}$, depending on the scenario. In line with our analysis plan, AFs and attributable/excess deaths are reported as point estimates, with statistical uncertainty conveyed by the confidence intervals of the underlying Cox model hazard ratios rather than by separate simulation-based confidence intervals for the PIFs. The main results for PIFs are illustrated in Figure~3 of the main manuscript.

**6.5 Proteomic mediation analysis**

For the biological extension, a two-stage mediation analysis was conducted in the proteomic subcohort to identify circulating biomarkers that might mediate associations between habitual UV exposure and cause-specific mortality from cardiovascular disease and cancer. This analysis used the same mortality outcomes, Sun-BEEM exposure parameterisation, and covariate set as the main multivariable Cox mortality models, and was motivated by prior work on UV-mediated immunomodulation and systemic disease [3].

In the first stage, for each biomarker we fitted two Cox proportional hazards models for the outcome in the proteomic subcohort: a model including Sun-BEEM and all covariates (total effect model) and a model additionally including the biomarker (direct effect model). Sun-BEEM was entered as a three-level categorical variable (low, medium, high), with low (score 0–1) as the reference category. Biomarkers were analysed on the NPX (log₂) scale. For each biomarker–outcome pair, the difference between the Sun-BEEM coefficients on the log-HR scale from the total and direct models was interpreted as the indirect (mediated) effect, and the proportion mediated was calculated as (total effect − direct effect) / total effect [42] [43]. Ninety-five per cent confidence intervals for the indirect effect were obtained using non-parametric bootstrap resampling with 1 000 iterations. Biomarkers with sparse data (fewer than 100 participants with valid biomarker measurements) or unstable bootstrap estimates (fewer than 10 successful bootstrap replications because of model non-convergence) were excluded.

In the second stage, associations between Sun-BEEM and each biomarker were examined using multivariable linear regression, with NPX as the outcome and Sun-BEEM coded as an ordinal variable. Models were adjusted for the same covariates as the main mortality model (age group, sex, BMI category, Townsend deprivation quintile, educational attainment, smoking status, physical activity category, sleep score, and alcohol intake). Analyses were conducted in the proteomic subcohort after excluding observations with missing biomarker, exposure, or covariate data; biomarkers with no variation in NPX values or with insufficient variation in Sun-BEEM (fewer than two exposure levels represented) were excluded.

For each biomarker we extracted the regression coefficient and p value for the Sun-BEEM trend term. To account for multiple testing across the panel of biomarkers, p values were adjusted using the Benjamini–Hochberg false discovery rate (FDR) procedure [44]. Biomarkers were classified as Sun-BEEM–responsive if the FDR-adjusted p value for the trend term was <0.05, with the direction of association defined by the sign of the regression coefficient (upregulated if the coefficient was positive, downregulated if negative).

Biomarkers were considered putative mediators if they met both of the following criteria: first, the bootstrapped 95% confidence interval for the indirect effect (from the first stage) excluded zero; and second, the FDR-adjusted p value for the Sun-BEEM–biomarker association (from the second stage) was <0.05, with a direction of effect consistent with the hypothesised mediation pathway. All statistical tests were two-sided. Biomarkers that fulfilled these criteria were then grouped into mechanistic “axes” (for example, immunoregulatory, mucosal–barrier/innate, cardiorenal–neuroendocrine) based on known biology and prior literature on UV-responsive pathways and cardiovascular or cancer risk, as detailed in Section 7 and Supplementary Tables S9–S11.

**6.6 Sensitivity and stratified analyses**

We first examined the influence of different covariate sets using nested stepwise Cox models with progressively extended adjustment, for both incidence and mortality outcomes. The resulting hazard ratios for Sun-BEEM categories and tests for trend are shown as forest plots in Supplementary Figures S1 (incident outcomes) and S2 (mortality outcomes.)

We then conducted pre-specified lagged (landmark) analyses for mortality outcomes, repeating Cox models after excluding deaths occurring within the first two years and, separately, within the first five years of follow-up, to assess potential reverse causation (for example, participants with pre-existing serious illness who might have reduced their UV exposure)[45]. Results of these analyses are shown as forest plots in Supplementary Figures S3 and S4 and were broadly consistent with the main analyses.

For stratified analyses by sex and age, we repeated fully adjusted Cox models stratified by sex (female, male) and by age (<60 vs ≥60 years at baseline), omitting the stratifying variable from the covariate set. Stratified HRs for Sun-BEEM categories and tests for trend were visualised as forest plots in Supplementary Figures S5 and S6; separate numerical tables were not provided because the plotted estimates were sufficient to summarise the patterns.

**7. Sun-BEEM–responsive biomarkers and biological axes**

Putative mediating proteins identified in the proteomic analyses were grouped into three broad biological axes. The immunoregulatory axis includes IL-22 and related mediators, reflecting T-cell and cytokine-mediated immune regulation with context-dependent roles in atherogenesis, hypertension, and tumour biology. IL-22 has been implicated in vascular dysfunction, blood pressure elevation, and endothelial injury in experimental models[46], and exhibits pro-tumour or anti-tumour activity depending on tissue context and microenvironment in colorectal and breast cancer [47] [48].

The mucosal–barrier/innate axis includes matrix metalloproteinase-7 (MMP-7), polymeric immunoglobulin receptor (PIGR), and related molecules involved in epithelial barrier integrity, innate defence, and extracellular matrix remodelling. Elevated circulating MMP-7 has been associated with plaque vulnerability and future cardiovascular events in carotid and peripheral arterial disease cohorts [49] [50]. PIGR is differentially expressed in gastrointestinal and respiratory tract cancers and has been linked to tumour differentiation and prognosis [51] [52].

The cardiorenal–neuroendocrine axis includes fibroblast growth factor 23 (FGF23) and renin–angiotensin system components, integrating UV-related effects on mineral metabolism, vascular phenotype, and neurohormonal regulation.Higher circulating FGF23 is consistently associated with increased cardiovascular events and mortality across multiple cohorts [53] [54].

Additional proteins, such as CD302 and LGALS4 (galectin-4), with reported links to immune cell trafficking, mucosal immunity, galectin biology, and tumour immune evasion, were also mapped onto these axes based on existing evidence.Higher expression of CD302 has been associated with poorer prognosis in multiple myeloma[55],while galectin-4 can act as a tumour suppressor in colorectal cancer[56], regulate intestinal T-cell apoptosis and inflammation [57], and, when produced extracellularly, promote T-cell apoptosis and immune evasion in pancreatic cancer[58]. A detailed summary of the direction of UV association, prior experimental or observational UV evidence, and published cardiovascular or cancer associations is provided in Supplementary Table S11.

**8. Overview of supplementary tables and figures**

**Supplementary tables**

- **Supplementary Table S1.** UV-related questionnaire items and coding for Sun-BEEM behavioural components.
- **Supplementary Table S2.** Environmental component used to construct the Sun-BEEM score (ambient shortwave radiation).
- **Supplementary Table S3.** UK Biobank covariates used in the main models and corresponding data-field identifiers.
- **Supplementary Table S4.** Mortality and incident outcomes: UK Biobank data sources, coding definitions and censoring dates.
- **Supplementary Table S5.** Serum 25-hydroxyvitamin D concentrations by binary Sun-BEEM components and categories.
- **Supplementary Table S6.** Childhood sunburn counts by binary Sun-BEEM components and categories.
- **Supplementary Table S9.** Top candidate protein mediators of the association between Sun-BEEM and cardiovascular mortality.
- **Supplementary Table S10.** Top candidate protein mediators of the association between Sun-BEEM and cancer mortality.
- **Supplementary Table S11.** Sun-BEEM–responsive biomarkers by mechanistic axis, reported UV associations, and prior links to cardiovascular and cancer risk.

**Supplementary figures**

- **Supplementary Figure S1.** Stepwise Cox proportional hazards models for incident outcomes according to Sun-BEEM category (nested adjustment from crude to fully adjusted models).
- **Supplementary Figure S2.** Stepwise Cox proportional hazards models for mortality outcomes according to Sun-BEEM category.
- **Supplementary Figure S3.** Landmark analyses: adjusted hazard ratios for mortality outcomes excluding deaths within the first 2 years of follow-up.
- **Supplementary Figure S4.** Landmark analyses: adjusted hazard ratios for mortality outcomes excluding deaths within the first 5 years of follow-up.
- **Supplementary Figure S5.** Sex-specific hazard ratios for mortality outcomes by Sun-BEEM exposure group (female and male strata).
- **Supplementary Figure S6.** Age-specific hazard ratios for mortality outcomes by Sun-BEEM exposure group (<60 and ≥60 years at baseline).

**Supplementary tables**

**Supplementary Table S1. UV-related questionnaire items and coding for Sun-BEEM behavioural components**

| **Sun-BEEM behavioural component** | **Baseline questionnaire item(s)*** | **Original response options (as used in this analysis)** | **Coding for Sun-BEEM component** |
| --- | --- | --- | --- |
| **Average time spent outdoors** | “On a typical day in summer, how many hours do you spend outdoors?” and “On a typical day in winter, how many hours do you spend outdoors?” | Participants reported hours per day (numeric). For each participant, summer and winter values were averaged to obtain a mean daily outdoor time. | Mean daily outdoor time <4 h/day → coded as 0 (lower behavioural UV exposure); mean daily outdoor time ≥4 h/day → coded as 1 (higher behavioural UV exposure). |
| **Solarium / sunlamp use** | “About how often do you use a solarium or sunlamp?” | Frequency of solarium or sunlamp use over the past year. Responses were collapsed to: ≥1 time per year vs <1 time per year or never. “Do not know” / “Prefer not to answer” were set to missing and excluded from Sun-BEEM construction. | <1 time/year or never → coded as 0 (no/low artificial UV use); ≥1 time/year → coded as 1 (artificial UV use). |
| **Sun/UV protection use** | “When you are outdoors in the summer, how often do you use sun/UV protection (such as sunscreen or protective clothing)?” | Responses were collapsed into two groups: “never/rarely” and “do not go out in sunshine” vs “sometimes”, “most of the time”, or “always”. | “Never/rarely” or “do not go out in sunshine” → coded as 0 (low behavioural UV exposure); “sometimes/most of the time/always” → coded as 1 (higher behavioural UV exposure prompting protection). |

**Supplementary Table S2. Environmental component used to construct the Sun-Behavioural and Environmental Exposure Model (Sun-BEEM) score**

| **Sun-BEEM environmental component** | **Data source / variable** | **Derivation and processing** | **Coding for Sun-BEEM component** |
| --- | --- | --- | --- |
| **Ambient short-wave radiation (SWR)** | Annual mean downward short-wave radiation (kJ/m²) at baseline residential location, derived from satellite-based products (e.g. MODIS Terra/Aqua–based land surface shortwave radiation) for the year 2008. | UK Biobank provided residential coordinates using the Ordnance Survey National Grid (1 km × 1 km). These were converted to latitude–longitude and spatially linked to the nearest grid cell in the short-wave radiation product. Monthly mean SWR values for 2008 were averaged to obtain an annual mean SWR (kJ/m²) for each grid cell. An a priori cut-off corresponding approximately to the median SWR at a centrally located assessment centre (Nottingham) was used to dichotomise exposure into “low” and “high” ambient solar radiation. | Annual mean SWR below the pre-defined cut-off → coded as 0 (lower ambient UV exposure); annual mean SWR at or above the cut-off → coded as 1 (higher ambient UV exposure). |

**Supplementary Table S3. UK Biobank covariates used in the main models and corresponding data-field identifiers**

| **Covariate** | **UK Biobank field ID(s)** | **Description / source** |
| --- | --- | --- |
| Age at recruitment | 21022 | Age at recruitment (years) |
| Sex | 31 | Sex (from central registry at recruitment) |
| Body mass index (BMI) | 21001 | Body mass index (kg/m²), derived from measured height and weight |
| Townsend deprivation index | 22189 | Townsend deprivation index at recruitment (area-level socioeconomic deprivation) |
| Educational attainment | 6138 | Qualifications (highest educational attainment) |
| Smoking status | 20116 | Smoking status (never/previous/current) |
| Alcohol drinker status | 20117 | Alcohol drinker status (never/previous/current) |
| Alcohol intake frequency | 1558 | Alcohol intake frequency (“About how often do you drink alcohol?”) |
| Total physical activity (MET-min/week) | 22040 | Total MET-minutes/week (walking, moderate and vigorous activity) |
| Chronotype | 1180 | Morning/evening person (chronotype) |
| Sleep duration | 1160 | Usual sleep duration (hours per 24 h period) |
| Sleeplessness / insomnia | 1200 | Sleeplessness / insomnia |
| Snoring | 1210 | Snoring |
| Daytime dozing / sleepiness | 1220 | Daytime dozing / sleeping |

**Supplementary Table S4. Mortality and incident outcomes: UK Biobank data fields and coding definitions**

| **Outcome** | **Type** | **Data source** | **UK Biobank field ID(s)** | **Codes used (underlying cause / main diagnosis)** |
| --- | --- | --- | --- | --- |
| All-cause mortality | Mortality | National death registry linkage | 40000 (date of death); 40001 (underlying cause of death); 40002 (contributory cause of death; not used in main analyses) | Any underlying cause of death |
| Cardiovascular disease (CVD) mortality (narrow) | Mortality | National death registry linkage | 40000; 40001 | I21–I25, I60–I64 (underlying cause of death) |
| Cancer mortality | Mortality | National death registry linkage | 40000; 40001 | C00–C97, D37–D48 (underlying cause of death) |
| Skin cancer mortality | Mortality | National death registry linkage | 40000; 40001 | C43–C44 (underlying cause of death) |
| Melanoma skin cancer mortality | Mortality | National death registry linkage | 40000; 40001 | C43 (underlying cause of death) |
| Non-melanoma skin cancer mortality | Mortality | National death registry linkage | 40000; 40001 | C44 (underlying cause of death) |
| Non-skin cancer mortality | Mortality | National death registry linkage | 40000; 40001 | C00–C97, D37–D48, excluding C43 and C44 (underlying cause of death) |
| Incident CVD (narrow: ischaemic heart disease and stroke) | Incidence | Hospital inpatient records (HES and Scottish equivalents) | 41270 (hospital inpatient diagnoses, ICD-10); 41271 (hospital inpatient diagnoses, ICD-9); 41280 (date of first in-patient diagnosis, ICD-10); 41281 (date of first in-patient diagnosis, ICD-9) | I21–I25, I60–I64 (any listed diagnosis in hospital inpatient records) |
| Incident cancer (all sites) | Incidence | National cancer registry linkage | 40005 (date of cancer diagnosis); 40006 (cancer type, ICD-10) | C00–C97, D37–D48 (first primary cancer diagnosis) |
| Incident melanoma skin cancer | Incidence | National cancer registry linkage | 40005; 40006 | C43 (first melanoma diagnosis) |
| Incident non-melanoma skin cancer | Incidence | National cancer registry linkage | 40005; 40006 | C44 (first keratinocyte carcinoma diagnosis) |
| Incident non-skin cancer | Incidence | National cancer registry linkage | 40005; 40006 | C00–C97, D37–D48, excluding C43 and C44 (first non-skin cancer diagnosis) |

**Supplementary Table S5. Serum 25-hydroxyvitamin D concentrations by binary Sun-BEEM component**

| **Component** | **Exposure Category** | **Mean 25(OH)D (nmol/L)** | **SD** | **p-value** |
| --- | --- | --- | --- | --- |
| Solarium use | No | 49.10 | 20.53 | <0.0001 |
|  | Yes | 65.07 | 23.62 |  |
| Residential SWR | Low (<9516 kJ/m²) | 48.94 | 21.14 | <0.0001 |
|  | High (≥9516 kJ/m²) | 51.40 | 20.55 |  |
| UV protection use | Rare/none | 45.29 | 21.19 | <0.0001 |
|  | Sometimes/often/always | 50.28 | 20.88 |  |
| Outdoor time | <4 hours/day | 48.66 | 20.69 | <0.0001 |
|  | ≥4 hours/day | 53.40 | 21.35 |  |

**Supplementary Table S6. Childhood sunburn counts by binary Sun-BEEM component**

| **Component** | **Exposure Category** | **Mean childhood sunburns** | **SD** | **p-value** |
| --- | --- | --- | --- | --- |
| Solarium use | No | 1.74 | 5.16 | <0.0001 |
|  | Yes | 1.50 | 2.81 |  |
| Residential SWR | Low (<9516 kJ/m²) | 1.54 | 4.70 | <0.0001 |
|  | High (≥9516 kJ/m²) | 2.06 | 5.64 |  |
| UV protection use | Rare/none | 1.49 | 3.92 | <0.0001 |
|  | Sometimes/often/always | 1.75 | 5.17 |  |
| Outdoor time | <4 hours/day | 1.79 | 4.65 | <0.0001 |
|  | ≥4 hours/day | 1.54 | 6.17 |  |

**Supplementary Table S7. Top candidate protein mediators of the association between Sun-BEEM and cardiovascular mortality**

| **Protein** | **β for Sun-BEEM (per category)*** | **Proportion of UV–CVD mortality association mediated** |
| --- | --- | --- |
| CD302 | −0.0092 | 0.18 (18.0%) |
| FGF23 | −0.0177 | 0.12 (12.3%) |
| LGALS4 | −0.0194 | 0.11 (11.1%) |
| REN | −0.0267 | 0.11 (10.9%) |
| CHGA | −0.0365 | 0.10 (10.2%) |
| FABP4 | −0.0134 | 0.10 (9.9%) |
| PIGR | −0.0142 | 0.10 (9.9%) |
| BGLAP | −0.0654 | 0.10 (9.8%) |
| NPDC1 | −0.0090 | 0.09 (9.2%) |
| NHLRC3 | −0.0179 | 0.09 (8.9%) |
| REG3A | −0.0226 | 0.08 (8.4%) |
| ADM | −0.0068 | 0.08 (8.4%) |
| GAST | −0.0394 | 0.08 (8.2%) |
| MMP7 | −0.0239 | 0.08 (7.8%) |
| IL22 | −0.0225 | 0.08 (7.7%) |

**Supplementary Table S8. Top candidate protein mediators of the association between Sun-BEEM and cancer mortality**

| **Protein** | **β for Sun-BEEM (per category)*** | **Proportion of UV–cancer mortality association mediated** |
| --- | --- | --- |
| PIGR | −0.0142 | 0.12 (11.9%) |
| NHLRC3 | −0.0179 | 0.12 (11.5%) |
| WFDC2 | −0.0126 | 0.11 (11.0%) |
| MMP7 | −0.0239 | 0.09 (9.0%) |
| KIT | 0.0266 | 0.09 (8.9%) |
| LGALS4 | −0.0194 | 0.09 (8.8%) |
| IL6 | −0.0334 | 0.09 (8.6%) |
| SIGLEC1 | −0.0132 | 0.08 (8.2%) |
| CD302 | −0.0092 | 0.08 (8.1%) |
| BST2 | −0.0221 | 0.08 (8.0%) |
| ICAM1 | −0.0070 | 0.08 (7.9%) |
| IL22 | −0.0225 | 0.08 (7.8%) |
| ITGA11 | 0.0187 | 0.07 (7.3%) |
| REN | −0.0267 | 0.07 (6.9%) |
| FGF23 | −0.0177 | 0.07 (6.9%) |

*Negative β coefficients indicate lower circulating protein concentrations at higher habitual Sun-BEEM exposure; positive β coefficients indicate higher concentrations.

**Supplementary Table S9. Sun-BEEM–responsive biomarkers by axis, reported UV-related evidence, and prior links to cardiovascular and cancer risk**

Arrows (↑/↓) indicate the direction reported in cited literature; “No” denotes that we did not identify direct experimental UV radiation evidence for that biomarker, and the cardiovascular disease (CVD)/cancer column summarises previously published associations and does not reflect the present mediation analysis.

| **Biomarker** | **Axis (as used in main text)** | **Evidence for association with UV exposure** | **Evidence for association with cardiovascular disease / cancer (prior literature)** |
| --- | --- | --- | --- |
| **IL-22** | Immunoregulatory | Experimental and clinical photobiology studies indicate that UV radiation and UV-based phototherapy modulate Th17/Th22-type cytokines, including IL-22, in skin and systemic immune responses.[59] [3] | IL-22 has been linked to vascular inflammation, endothelial dysfunction, and hypertension[46],and exerts context-dependent tumour-promoting effects in colorectal and breast cancer models.[47] [60] |
| **NHLRC3** | Immunoregulatory | No direct experimental UV radiation data identified. | No robust published evidence directly linking circulating or tissue NHLRC3 levels to cardiovascular events or major cancers was identified in our focused search. |
| **CD302** | Mucosal–barrier / innate | No direct experimental UV radiation data identified. | CD302 has disease-specific prognostic implications, functioning as an adverse marker in chronic myeloid leukaemia treated with tyrosine kinase inhibitors and in low-grade glioma[61] [62], whereas higher CD302 expression is associated with more favourable outcomes in multiple myeloma and lung adenocarcinoma[63] [64]. |
| **PIGR** | Mucosal–barrier / innate | No direct experimental UV radiation data identified. | PIGR shows context-dependent roles across epithelial cancers: loss or reduced expression has been linked to more aggressive colorectal cancer behaviour[51] ,whereas overexpression has been associated with adverse outcomes in nasopharyngeal carcinoma and other upper gastrointestinal adenocarcinomas[52] [65] |
| **LGALS4 (Galectin-4)** | Mucosal–barrier / innate | No direct UV radiation data identified. | Galectin-4 can act as a tumour suppressor in colorectal cancer[56],but extracellular galectin-4 also induces T-cell apoptosis and blocks antitumour immunity in pancreatic ductal adenocarcinoma and functions as a protumourigenic immunomodulator in other gastrointestinal malignancies, highlighting a context-dependent role in cancer progression[58] [66] |
| **MMP-7** | Mucosal–barrier / matrix | No direct human systemic UV data identified; matrix metalloproteinases, including MMP-7, are involved in UV-driven skin remodelling and photoageing pathways summarised in photoimmunology reviews[67]. | Elevated circulating MMP-7 has been associated with higher risk of recurrent vascular events after carotid endarterectomy and with major adverse cardiovascular events in peripheral artery disease, suggesting a link between MMP-7 and atherosclerotic disease progression [68] [69]. |
| **FGF23** | Cardiorenal–neuroendocrine | FGF23 lies on mineral metabolism and vitamin D–related pathways that can be influenced by UV exposure and UV-based phototherapy, although direct long-term UV effects on circulating FGF23 remain incompletely characterised.[70] | Higher FGF23 concentrations have consistently been associated with increased CVD events and all-cause mortality in chronic kidney disease and general-population cohorts [53] [71]. |
| **REN (Renin / plasma renin activity)** | Cardiorenal–neuroendocrine | No direct experimental UV radiation evidence identified for REN/plasma renin activity. | Higher plasma renin activity is widely reported to predict cardiovascular events and mortality in hypertensive and high-risk cohorts. |

**Supplementary figures**

**Supplementary Figure S1. Stepwise Cox proportional hazards models for incident outcomes according to Sun-BEEM category**

Hazard ratios (95% CIs) for medium and high Sun-BEEM exposure versus low exposure are shown from stepwise Cox models for cardiovascular disease, all cancer, non-skin cancer, melanoma, and other skin cancer incidence. Models 1–10 add covariates sequentially; circles indicate medium versus low, squares high versus low, and the vertical dashed line marks the null (HR=1).

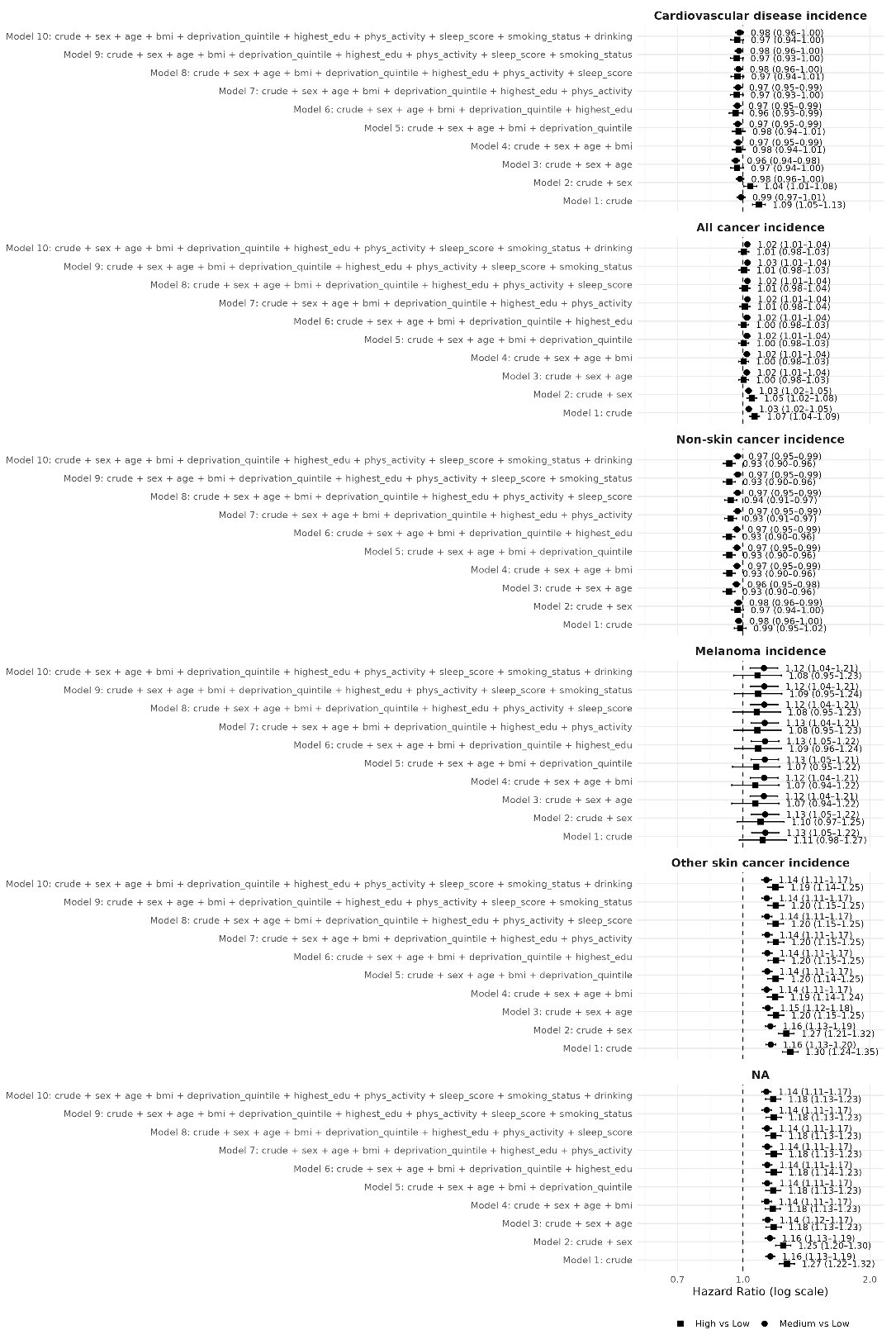

**Supplementary Figure S2. Stepwise Cox proportional hazards models for mortality outcomes according to Sun-BEEM category**

Hazard ratios (95% CIs) for medium and high Sun-BEEM exposure versus low exposure are shown for all-cause, cardiovascular disease, cancer, cancer excluding skin cancer, melanoma, and other skin cancer mortality. Models 1–10 add covariates sequentially. Circles indicate medium versus low exposure, squares high versus low; the vertical dashed line marks the null (HR=1).

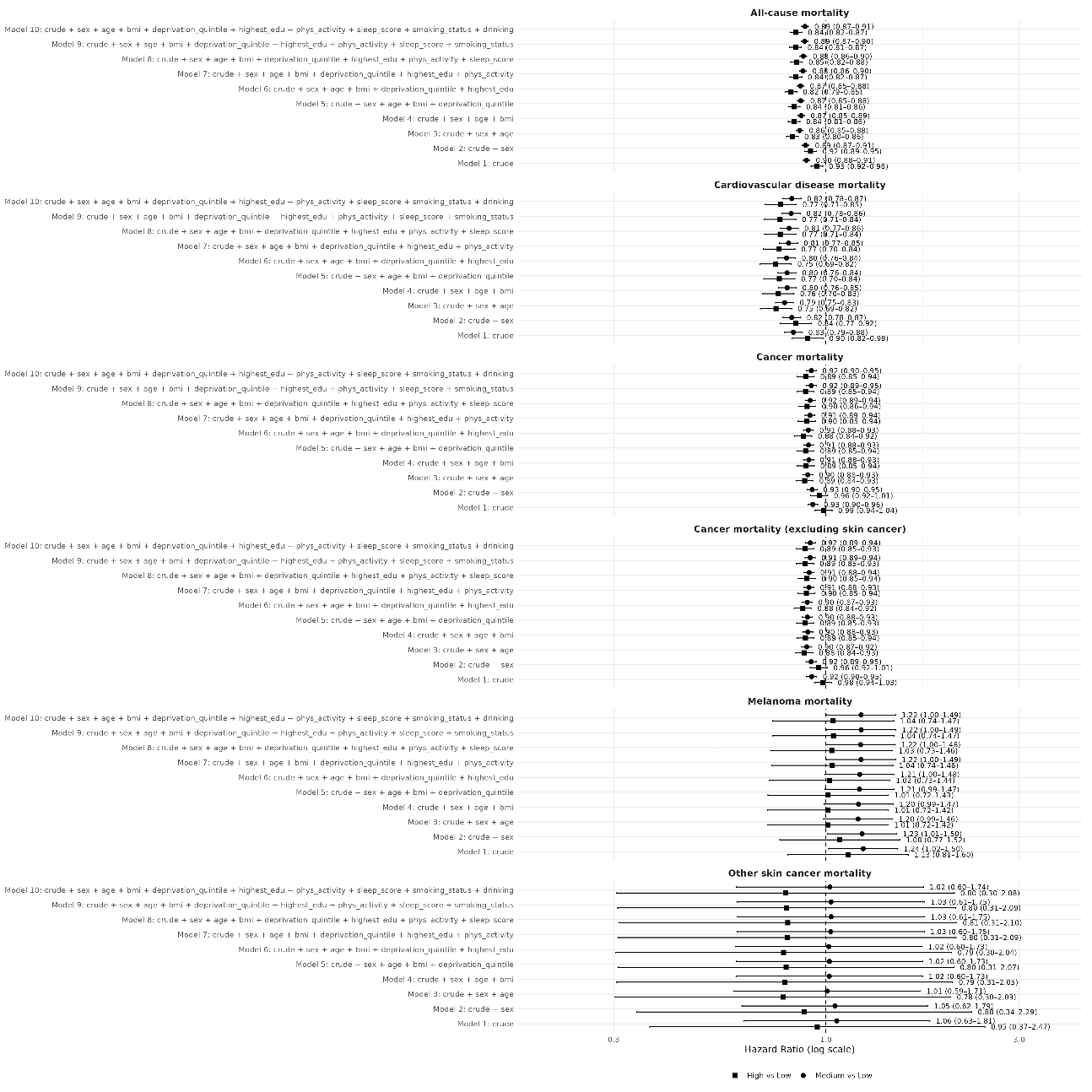

**Supplementary Figure S3. Landmark analyses: adjusted hazard ratios for mortality outcomes excluding deaths within the first 2 years of follow-up**
Hazard ratios for mortality outcomes by Sun-BEEM exposure category are estimated from Cox proportional hazards models excluding participants who died within the first 2 years of follow-up. Models are adjusted for all covariates in the main model. Circles represent medium versus low UV exposure and triangles represent high versus low UV exposure; horizontal lines show 95% confidence intervals. The x-axis is on a logarithmic scale and the dashed vertical line indicates a hazard ratio of 1.0.

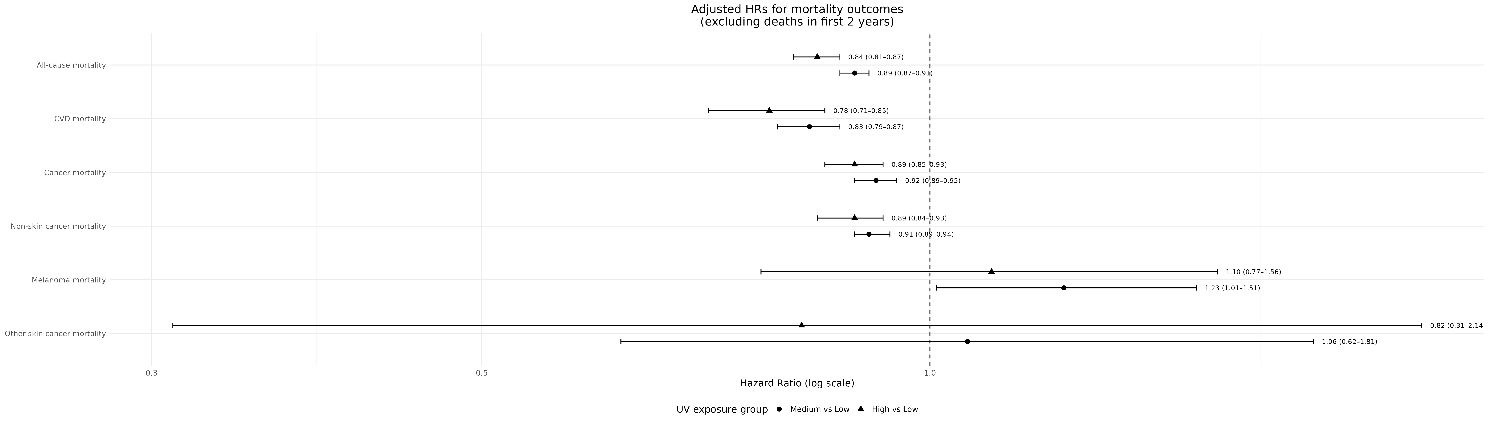

**Supplementary Figure S4. Landmark analyses: adjusted hazard ratios for mortality outcomes excluding deaths within the first 5 years of follow-up**
Hazard ratios for mortality outcomes by Sun-BEEM exposure category are estimated from Cox proportional hazards models excluding participants who died within the first 5 years of follow-up. Models are adjusted for all covariates in the main model. Circles represent medium versus low UV exposure and triangles represent high versus low UV exposure; horizontal lines show 95% confidence intervals. The x-axis is on a logarithmic scale and the dashed vertical line indicates a hazard ratio of 1.0.

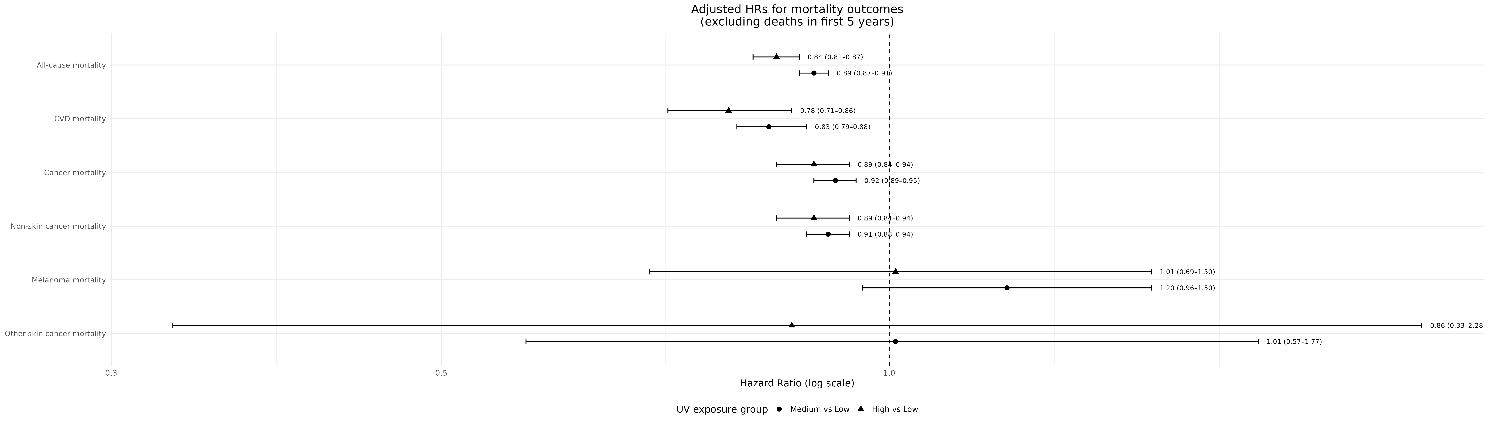

**Supplementary Figure S5. Sex-specific hazard ratios for mortality outcomes by Sun-BEEM exposure group**
Hazard ratios are from multivariable Cox proportional hazards models adjusted for all covariates in the main model and are shown separately for women and men (distinguished by colour). Circles represent medium versus low UV exposure and triangles represent high versus low UV exposure; horizontal lines show 95% confidence intervals. The x-axis is on a logarithmic scale and the dashed vertical line indicates a hazard ratio of 1.0.

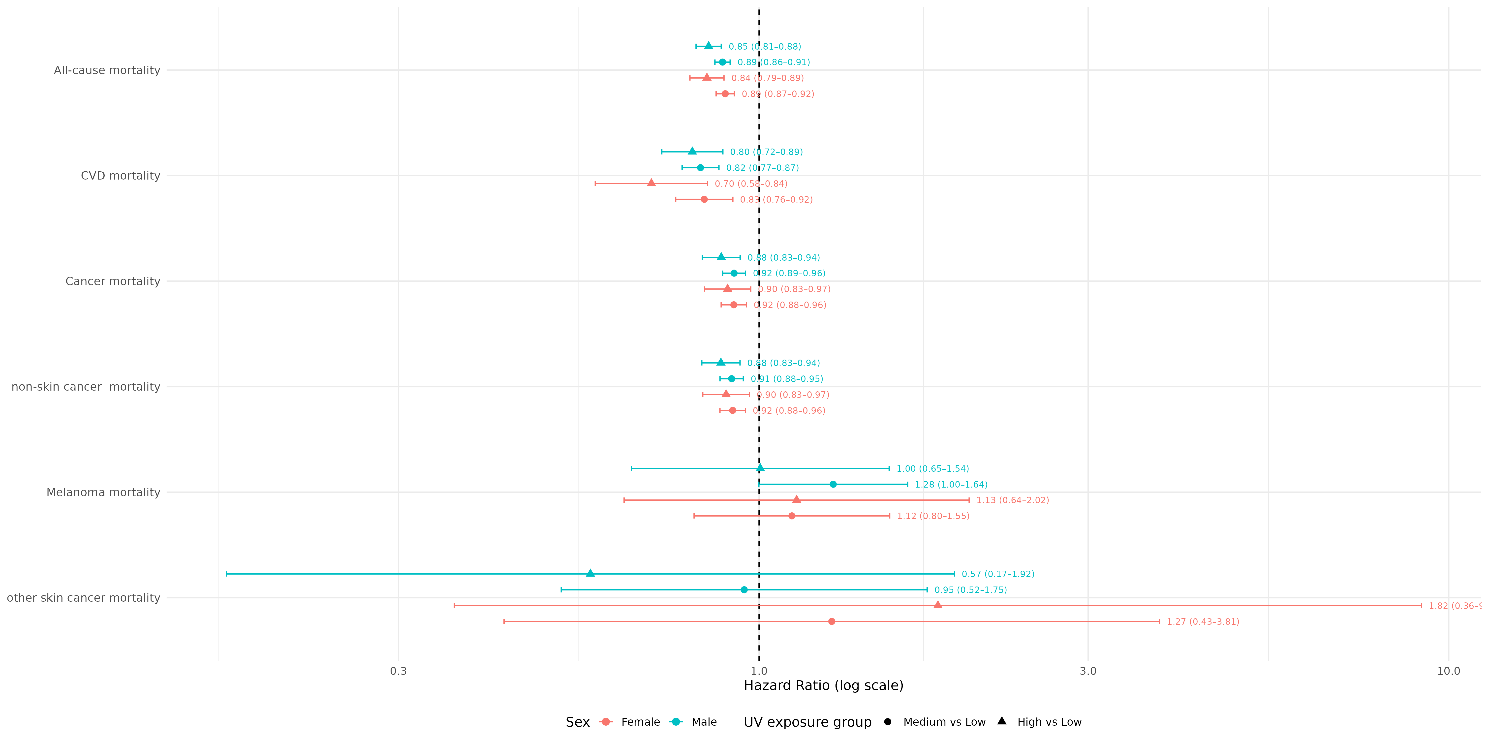

**Supplementary Figure S6. Age-specific hazard ratios for mortality outcomes by Sun-BEEM exposure group**
Hazard ratios are from multivariable Cox proportional hazards models adjusted for all covariates in the main model and are shown separately for participants aged <60 years and ≥60 years (distinguished by colour). Circles represent medium versus low UV exposure and triangles represent high versus low UV exposure; horizontal lines show 95% confidence intervals. The x-axis is on a logarithmic scale and the dashed vertical line indicates a hazard ratio of 1.0.

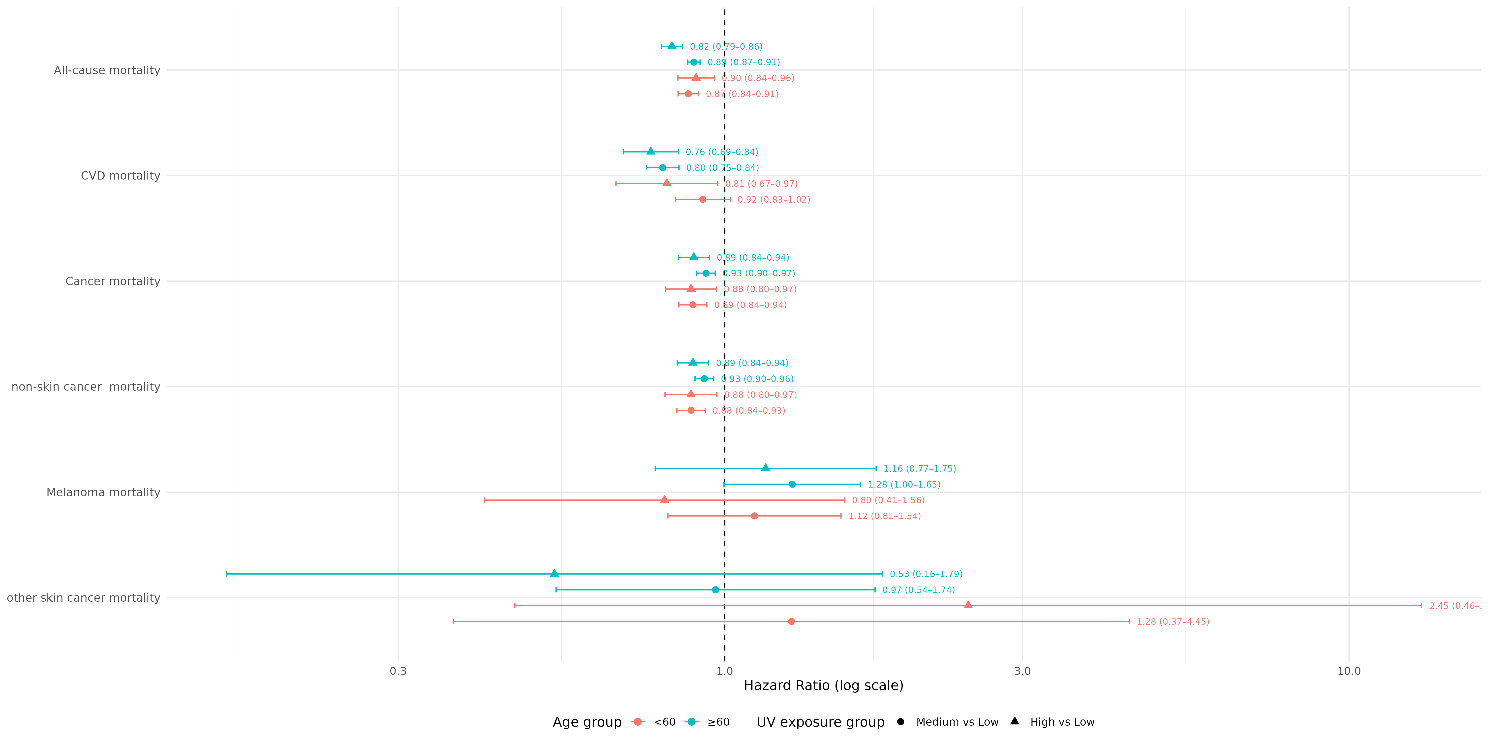

*Abbreviations for figures: HR, hazard ratio; CI, confidence interval; CVD, cardiovascular disease.

**Supplementary references**

1	Sudlow C, Gallacher J, Allen N. UK Biobank: an open access resource for identifying the causes of a wide range of complex diseases of middle and old age. *PLoS Med*. 2015;12:e1001779.

2	Assarsson E, Lundberg M, Holmquist G. Homogenous 96-plex PEA immunoassay exhibiting high sensitivity, specificity, and excellent scalability. *PLoS One*. 2014;9:e95192.

3	Hart PH, Norval M, Byrne SN, *et al.* Exposure to ultraviolet radiation in the modulation of human diseases. *Annu Rev Pathol*. 2019;14:55–81.

4	Shentu Y, Xie M. A note on dichotomization of continuous response variable in the presence of contamination and model misspecification. *Stat Med*. 2010;29:2200–14. doi: 10.1002/sim.3966

5	Li Y, Schoufour J, Wang DD. Healthy lifestyle and life expectancy free of cancer, cardiovascular disease, and type 2 diabetes: prospective cohort study. *BMJ*. 2020;368:l6669.

6	Chudasama YV, Khunti K, Gillies CL. Healthy lifestyle and life expectancy in people with multimorbidity in the UK Biobank: a longitudinal cohort study. *PLoS Med*. 2020;17:e1003332.

7	Nylund-Gibson K, Garber AC, Carter DB, *et al.* Ten frequently asked questions about latent transition analysis. *Psychol Methods*. 2023;28:284–300. doi: 10.1037/met0000486

8	Nylund KL, Asparouhov T, Muthén BO. Deciding on the Number of Classes in Latent Class Analysis and Growth Mixture Modeling: A Monte Carlo Simulation Study. *Structural Equation Modeling: A Multidisciplinary Journal*. 2007;14:535–69. doi: 10.1080/10705510701575396

9	Rousseeuw PJ. Silhouettes: A graphical aid to the interpretation and validation of cluster analysis. *Journal of Computational and Applied Mathematics*. 1987;20:53–65. doi: 10.1016/0377-0427(87)90125-7

10	NASA Earthdata (LP DAAC). *MODIS/Terra+Aqua Surface Radiation Daily/3-Hour L3 Global 1km SIN Grid (MCD18A1)* (Version 6.1/6.2). [https://www.earthdata.nasa.gov/data/catalog/lpcloud-mcd18a1-061](https://www.earthdata.nasa.gov/data/catalog/lpcloud-mcd18a1-061?utm_source=chatgpt.com) (accessed 18 Dec 2025).

11	Diffey BL. An overview analysis of the time people spend outdoors. *Br J Dermatol*. 2011;164:848–55.

12	Baczynska KA, Khazova M, O’Hagan JB. Sun exposure of indoor workers in the UK: survey on the time spent outdoors. *Photochem Photobiol Sci*. 2019;18:120–8.

13	Soueid L, Triguero-Mas M, Dalmau A. Estimating personal solar ultraviolet radiation exposure through time spent outdoors, ambient levels and modelling approaches. *Br J Dermatol*. 2022;186:266–73.

14	Boniol M, Autier P, Boyle P, *et al.* Cutaneous melanoma attributable to sunbed use: systematic review and meta-analysis. *BMJ*. 2012;345:e4757. doi: 10.1136/bmj.e4757

15	International Agency for Research on Cancer (IARC) Working Group on Risk of Skin Cancer and Exposure to Artificial Ultraviolet Light. *Exposure to artificial UV radiation and skin cancer*. IARC Working Group Reports, Vol 1. Lyon: International Agency for Research on Cancer; 2006.

16	Wehner MR, Shive ML, Chren M-M, *et al.* Indoor tanning and non-melanoma skin cancer: systematic review and meta-analysis. *BMJ*. 2012;345:e5909. doi: 10.1136/bmj.e5909

17	Thieden E, Philipsen PA, Sandby-Møller J, *et al.* Sunscreen use related to UV exposure, age, sex, and occupation based on personal dosimeter readings and sun-exposure behavior diaries. *Arch Dermatol*. 2005;141:967–73. doi: 10.1001/archderm.141.8.967

18	Autier P, Doré JF, Négrier S. Sunscreen use and duration of sun exposure: a double-blind, randomized trial. *J Natl Cancer Inst*. 1999;91:1304–9.

19	International Agency for Research on Cancer. *Sunscreens*. IARC Handbook of Cancer Prevention, Vol 5. Lyon: IARC Press; 2001.

20	Holick MF. Vitamin D deficiency. *N Engl J Med*. 2007;357:266–81.

21	Armstrong BK, Kricker A. How much melanoma is caused by sun exposure? *Melanoma Res*. 1993;3:395–401.

22	Fry A, Littlejohns TJ, Sudlow C, et al.Comparison of sociodemographic and health-related characteristics of UK Biobank participants with the general population. *American Journal of Epidemiology.* 2017;186(9):1026–1034. doi：10.1093/aje/kwx246

23	Boakye D, Wyse CA, Morales-Celis CA, *et al.* Tobacco exposure and sleep disturbance in 498 208 UK Biobank participants. *J Public Health (Oxf)*. 2018;40:517–26. doi: 10.1093/pubmed/fdx102

24	UK Chief Medical Officers. *UK Chief Medical Officers’ low risk drinking guidelines*. London: Department of Health; 2016.

25	IPAQ Research Committee. Guidelines for data processing and analysis of the International Physical Activity Questionnaire (IPAQ). Revised November 2005. Available from: https://sites.google.com/site/theipaq/scoring-protocol (accessed 18 Dec 2025).

26	Chudasama YV, Khunti KK, Zaccardi F, *et al.* Physical activity, multimorbidity, and life expectancy: a UK Biobank longitudinal study. *BMC Med*. 2019;17:108. doi: 10.1186/s12916-019-1339-0

27	Fan M, Sun D, Zhou T, *et al.* Sleep patterns, genetic susceptibility, and incident cardiovascular disease: a prospective study of 385 292 UK biobank participants. *Eur Heart J*. 2020;41:1182–9. doi: 10.1093/eurheartj/ehz849

28	White IR, Royston P, Wood AM. Multiple imputation using chained equations: Issues and guidance for practice. *Stat Med*. 2011;30:377–99. doi: 10.1002/sim.4067

29	van Buuren S, Groothuis-Oudshoorn K. mice: multivariate imputation by chained equations in R. *J Stat Softw*. 2011;45:1–67.

30	Schnier C, et al. Identifying myocardial infarction and stroke in UK Biobank: phenotypes and algorithms for linked health data. *International Journal of Population Data Science.* 2017. doi:10.23889/ijpds.v1i1.358

31	Parra-Soto S, et al. Associations of six adiposity-related markers with incidence and mortality from 24 cancers—findings from the UK Biobank prospective cohort study. *BMC Medicine*, 2021;19:7 doi:10.1186/s12916-020-01848-8.

32	Elliott P, Peakman TC, UK Biobank. The UK Biobank sample handling and storage protocol for the collection, processing and archiving of human blood and urine. *Int J Epidemiol*. 2008;37:234–44. doi: 10.1093/ije/dym276

33	Wik L, Nordberg N, Broberg J, *et al.* Proximity Extension Assay in Combination with Next-Generation Sequencing for High-throughput Proteome-wide Analysis. *Mol Cell Proteomics*. 2021;20:100168. doi: 10.1016/j.mcpro.2021.100168

34	Sun BB, Chiou J, Traylor M, *et al.* Plasma proteomic associations with genetics and health in the UK Biobank. *Nature*. 2023;622:329–38. doi: 10.1038/s41586-023-06592-6

35 R Core Team. *R: a language and environment for statistical computing*. Vienna, Austria: R Foundation for Statistical Computing; 2024

36	Cox DR. Regression models and life-tables (with discussion). J R Stat Soc B 1972;34:187–220. doi:10.1111/j.2517-6161.1972.tb00899.x.

37	Therneau TM. *A package for survival analysis in R*. R package version 3.8-3. 2024.

38	Harrell FE. *Regression modeling strategies*. New York: Springer 2015.

39	Schoenfeld D. Partial residuals for the proportional hazards regression model. Biometrika 1982;69:239–41. doi:10.1093/biomet/69.1.239.

40	Keil AP, Edwards JK, Richardson DB, et al. The parametric g-formula for time-to-event data: intuition and a worked example. *Epidemiology.* 2014 Nov;25(6):889-97. doi: 10.1097/EDE.0000000000000160..

41	Mansournia MA, Altman DG. Population attributable fraction. *BMJ.* 2018;360:k757. doi:10.1136/bmj.k757.

42	VanderWeele TJ. Causal mediation analysis with survival data. *Epidemiology*. 2011;22:582–5. doi: 10.1097/EDE.0b013e31821db37e

43	VanderWeele TJ. Explanation in causal inference: developments in mediation and interaction. *Int J Epidemiol*. 2016;45:1904–8. doi: 10.1093/ije/dyw277

44	Benjamini Y, Hochberg Y. Controlling the False Discovery Rate: A Practical and Powerful Approach to Multiple Testing. *Journal of the Royal Statistical Society: Series B (Methodological)*. 1995;57:289–300. doi: 10.1111/j.2517-6161.1995.tb02031.x

45	Houwelingen H van, Putter H. *Dynamic Prediction in Clinical Survival Analysis*. Boca Raton: CRC Press 2011.

46	Ye J, Ji Q, Liu J, et al. Interleukin 22 promotes blood pressure elevation and endothelial dysfunction in angiotensin II-treated mice. *J Am Heart Assoc* 2017;6(10):e005875. doi:10.1161/JAHA.117.005875.

47	Kim K, Kim G, Kim JY. Interleukin-22 promotes breast cancer cell proliferation and migration. *Carcinogenesis*. 2014;35:1352–61.

48	Jiang R, Tan Z, Deng L, et al. Interleukin-22 is related to development of human colon cancer by activation of STAT3. BMC Cancer 2013;13:59. doi:10.1186/1471-2407-13-59

49	Abbas A, et al. Matrix metalloproteinase 7 is associated with symptomatic lesions and adverse events in patients with carotid atherosclerosis. *PLoS One* 2014;9(1):e84935.

50	Li B, Shaikh F, Younes H, et al. Matrix metalloproteinases 7 and 10 are prognostic biomarkers for systemic cardiovascular risk in individuals with peripheral artery disease. *Biomolecules* 2025;15(6):853. doi:10.3390/biom15060853.

51	Zhang D, Huang H, Zheng T. Polymeric immunoglobulin receptor suppresses colorectal cancer through the AKT–FOXO3/4 axis by downregulating LAMB3 expression. *Front Oncol* 2022;12:924988. doi:10.3389/fonc.2022.924988

52	Qi X, Li X, Sun X. Reduced expression of polymeric immunoglobulin receptor (PIGR) in nasopharyngeal carcinoma and its correlation with prognosis. *Tumour Biol* 2016;37:11099–104. doi:10.1007/s13277-016-4791-x

53	Lutsey PL, Alonso A, Selvin E. Fibroblast growth factor-23 and incident coronary heart disease, heart failure, and cardiovascular mortality: the ARIC study. *J Am Heart Assoc*. 2014;3:e000936.

54	Marthi A, Donovan K, Haynes R. Fibroblast growth factor-23 and risks of cardiovascular and noncardiovascular diseases: a meta-analysis. *J Am Soc Nephrol* 2018;29:2015–27. doi:10.1681/ASN.2017121334

55	Alaterre E, Raimbault S, Goldschmidt H, et al. CD24, CD27, CD36 and CD302 gene expression for outcome prediction in patients with multiple myeloma. *Oncotarget* 2017;8:98931–44. doi:10.18632/oncotarget.22131

56	Satelli A, Rao PS, Thirumala S, et al. Galectin-4 functions as a tumor suppressor of human colorectal cancer. *Int J Cancer* 2011;129:799–809. doi:10.1002/ijc.25750

57	Paclik D, Danese S, Berndt U, et al. Galectin-4 controls intestinal inflammation by selective regulation of peripheral and mucosal T cell apoptosis and cell cycle. *PLoS One* 2008;3(7):e2629. doi:10.1371/journal.pone.0002629

58	Lidström T, Cumming J, Gaur R, **et al**. Extracellular galectin 4 drives immune evasion and promotes T-cell apoptosis in pancreatic cancer. *Cancer Immunol Res* 2023;11(1):72–92. doi:10.1158/2326-6066.CIR-21-1088.

59	Vieyra-Garcia PA, Wolf P. A deep dive into UV-based phototherapy: Mechanisms of action and emerging molecular targets in inflammation and cancer. *Pharmacol Ther*. 2021;222:107784. doi: 10.1016/j.pharmthera.2020.107784

60	Kryczek I, Lin Y, Nagarsheth N, *et al.* IL-22(+)CD4(+) T cells promote colorectal cancer stemness via STAT3 transcription factor activation and induction of the methyltransferase DOT1L. *Immunity*. 2014;40:772–84. doi: 10.1016/j.immuni.2014.03.010

61	An Y, Wang Q, Zhang L, et al. OSlgg: an online prognostic biomarker analysis tool for low-grade glioma. Front Oncol 2020;10:1097. doi:10.3389/fonc.2020.01097

62	Kok CH, Irani Y, Clarson J, *et al.* CD302 predicts achievement of deep molecular response in patients with chronic myeloid leukemia treated with imatinib. *Blood Neoplasia*. 2024;1:100014. doi: 10.1016/j.bneo.2024.100014

63	Wang K, Wang Y, Lu T, *et al.* CD302 regulates the malignant phenotypes of lung adenocarcinoma as a tumor suppressor gene. *Front Oncol*. 2025;15:1601706. doi: 10.3389/fonc.2025.1601706

64	Alaterre E, Raimbault S, Goldschmidt H, *et al.* CD24, CD27, CD36 and CD302 gene expression for outcome prediction in patients with multiple myeloma. *Oncotarget*. 2017;8:98931–44. doi: 10.18632/oncotarget.22131

65	Fristedt R, Gaber A, Hedner C, *et al.* Expression and prognostic significance of the polymeric immunoglobulin receptor in esophageal and gastric adenocarcinoma. *J Transl Med*. 2014;12:83. doi: 10.1186/1479-5876-12-83

66	Vopálenská A, Balogová S, Hovorková M, *et al.* Targeting galectin-4 with glycoconjugates of varying architectures: a multivalency étude with accent on anti-tumor effect and protection against apoptosis. *Eur J Med Chem*. 2025;300:118149. doi: 10.1016/j.ejmech.2025.118149

67	Pittayapruek P, Meephansan J, Prapapan O, *et al.* Role of Matrix Metalloproteinases in Photoaging and Photocarcinogenesis. *Int J Mol Sci*. 2016;17:868. doi: 10.3390/ijms17060868

68	Moreno-Ajona D, Irimia P, Rodríguez JA, *et al.* Elevated circulating metalloproteinase 7 predicts recurrent cardiovascular events in patients with carotid stenosis: a prospective cohort study. *BMC Cardiovasc Disord*. 2020;20:93. doi: 10.1186/s12872-020-01387-3

69	Li B, Shaikh F, Younes H, *et al.* Matrix Metalloproteinases 7 and 10 Are Prognostic Biomarkers for Systemic Cardiovascular Risk in Individuals with Peripheral Artery Disease. *Biomolecules*. 2025;15:853. doi: 10.3390/biom15060853

70	Lee JH, Kim KM, Shin DY, *et al.* Ultraviolet B activated 1,25(OH)(2)D affects the level of fibroblast growth factor-23 in human. *Endocr J*. 2013;60:81–6. doi: 10.1507/endocrj.ej12-0199

71	Kendrick J, Cheung AK, Kaufman JS, *et al.* FGF-23 associates with death, cardiovascular events, and initiation of chronic dialysis. *J Am Soc Nephrol*. 2011;22:1913–22. doi: 10.1681/ASN.2010121224
